# Early-Life Sugar Restriction and Long-Term Risk of Cancer: A Natural Experiment Study in the UK

**DOI:** 10.1101/2025.11.22.25340786

**Authors:** Chen Zhu, Weilong Zhang

## Abstract

**Objective:** To examine whether early-life exposure to sugar rationing is associated with reduced risk of cancer and cardiometabolic disease in adulthood, leveraging a natural experiment created by the abrupt end of post-war sugar rationing in the United Kingdom on 26 September 1953.

**Design:** Natural experiment study using an event-study design based on variation in the duration of exposure to sugar rationing during the first 1000 days of life (from conception to age 2 years).

**Setting:** UK Biobank, a large prospective cohort recruited from across the UK.

**Participants:** ’64,761 UK Biobank participants born between 1951 and 1956, a window spanning the 1953 de-rationing event. Exposure to sugar restriction was determined by birth date relative to the policy change. Individuals with multiple births, adoption, birth outside the UK, or pre-existing disease were excluded.

**Main outcome measures:** Adult incidence of major cancers, digestive (liver, rectum), respiratory (lung), and hormone-sensitive (prostate, breast), estimated using Cox proportional hazards models. We also assessed long-term behavioural outcomes (dietary patterns) and biological markers (leukocyte telomere length; circulating Granzyme B).

**Results:** Longer exposure to sugar rationing during early life was associated with lower risk of multiple diseases in adulthood. Participants exposed in utero plus one to two years had lower incidence of digestive cancers (liver: hazard ratio 0.31, 95% confidence interval 0.18 to 0.49; rectum: 0.60, 0.51 to 0.69), respiratory cancer (lung: 0.59, 0.50 to 0.68), and hormone-sensitive cancers (prostate: 0.48, 0.43 to 0.55; breast: 0.64, 0.58 to 0.70).

**Mechanisms:** Two complementary mechanisms were identified: (1) a behavioural programming pathway, wherein early-life restriction led to a persistent hedonic shift resulting in lower sugar intake and healthier dietary habits five decades later; and (2) a biological imprinting pathway, evidenced by 0.05 SD longer leukocyte telomere length (*≈* 2.2 years less biological ageing) and lower circulating Granzyme B levels.

**Conclusion:** Exposure to sugar restriction during the first 1,000 days was associated with lower cancer and slower biological ageing, offering rare causal evidence that early-life nutrition can permanently shape disease susceptibility. During rationing, adults consumed about 40 g/day of sugar, well within WHO-recommended levels, whereas intake doubled to roughly 80 g/day once controls ended. This natural contrast shows that maintaining WHO-level sugar intake in early life can yield lasting health benefits. With current consumption far above recommended thresholds, the case for early-life sugar reduction is both urgent and highly consequential.

**What is already known on this topic:**

- Excess sugar intake is linked to metabolic and inflammatory pathways that increase the risk of chronic disease, but most human evidence is observational and potentially confounded.
- Prior studies have used the post-war sugar rationing period in the United Kingdom to identify long-term effects of early-life nutrition on metabolic and cardiovascular outcomes.
- No previous research has examined the causal impact of early-life sugar exposure on cancer incidence or biological ageing.

**What this study adds:**

- Leveraging the abrupt end of post-war sugar rationing in 1953 as a natural experiment, this study provides causal evidence that early-life sugar restriction reduces lifetime risk of cancer and cardiometabolic diseases.
- Early-life exposure to sugar rationing was associated with lower incidence of digestive (liver, rectum), respiratory (lung), and hormone-sensitive (prostate, breast) cancers.
- Two complementary mechanisms were identified: (1) a behavioural programming pathway, where early nutritional scarcity induced a persistent *hedonic shift* toward lower sugar intake and healthier diets; and (2) a biological imprinting pathway, reflected in longer leukocyte telomere length and lower Granzyme B levels, indicating slower cellular ageing and reduced inflammation.

## 1 Introduction

High sugar consumption is now a routine feature of diets across many countries, a significant public health concern highlighted in the United Nations’ 2030 Agenda for Sustainable Development to “ensure healthy lives and promote well-being for all at all ages” [1]. A growing body of evidence shows that sugar has addictive properties, with early exposure capable of altering neural reward pathways and shaping persistent taste preferences [2, 3]. Such patterns formed in childhood can reinforce long-term consumption habits and elevate lifetime health risks. Establishing whether early-life sugar intake contributes to chronic disease in adulthood is therefore an important policy priority.

The notion that “sugar feeds cancer” rests on well-established biological mechanisms. Elevated blood glucose levels induce insulin resistance, promoting tumour growth through enhanced glucose uptake and metabolic reprogramming of cancer cells [4]. High sugar intake also stimulates inflammatory cytokines and insulin-like growth factors (IGFs), which drive cell proliferation, inhibit apoptosis, and facilitate metastasis [5, 6]. Consistent with these pathways, several large prospective studies and meta-analyses report positive associations between high consumption of sugar-sweetened beverages or high-glycaemic-load diets and greater incidence of breast, colorectal, and pancreatic cancers [7–9].

Despite its biological plausibility, the empirical evidence for a causal relationship remains limited. Randomised controlled trials of long-term sugar exposure are ethically and practically infeasible, leaving a critical gap in causal inference.^1^ Most existing studies are observational and thus prone to omitted variable bias and reverse causality. Unobserved confounders, such as socioeconomic status, body mass index, and correlated dietary patterns, can distort estimated effects [7–9, 14]. The few quasi-causal studies available rely on Mendelian randomisation (MR) analyses. These show that genetically predicted glycaemic traits, such as elevated fasting insulin and glucose levels, are associated with higher risks of colorectal, endometrial, kidney, and breast cancers [15, 16], and that genetically proxied higher sugar intake may increase the risk of certain breast cancer subtypes [17]. However, such genetic instruments reflect metabolic genetic predispositions rather than individuals’ actual dietary behaviour.

A further limitation of the existing literature is its short temporal perspective. The health consequences of sugar exposure likely accumulate over decades, yet most studies focus on contemporary or short-term consumption, overlooking how nutritional conditions in early life might shape later disease susceptibility [18–20]. Evidence from developmental-origins research demonstrates that early nutritional environments, particularly during the first 1,000 days from conception to age two, exert lasting effects on metabolism, immunity, and chronic disease risk [21, 22]. However, no study has yet leveraged exogenous variation in early-life sugar availability to identify the long-term causal impact of reduced sugar exposure on cancer.

This study addresses these limitations by exploiting a unique natural experiment: the termination of sugar and sweets rationing in the United Kingdom in September 1953. The sugar-rationing policy, introduced in July 1942 as part of a broader fourteen-year wartime food-rationing programme, aimed to ensure equitable food distribution and prevent shortages during and after the Second World War [23]. During rationing, each person, including pregnant women and children aged two and above, received approximately 8 ounces (*≈* 225 g) of sugar per week and 12 ounces (*≈* 340 g) of sweets per month through a ration-book system; children under two years were not allocated sugar or sweets.

After rationing ended in September 1953, sugar consumption rose sharply. Drawing on the framework developed by Gracner and her colleagues [24, 25], we rely on their documented evidence that average daily adult sugar intake nearly doubled, from about 41 g in early 1953 to 80 g by mid-1954, while intake of other major foods and nutrients remained largely stable. This abrupt, sugar-specific shift created quasi-experimental variation in early-life exposure during a critical developmental window. Cohorts conceived just before September 1953 experienced restricted sugar access in utero and early childhood, whereas those conceived immediately afterward were exposed to sudden abundance. This design enables estimation of the causal effect of early-life sugar exposure on adult health outcomes, including cancer risk, while limiting confounding from lifestyle or socioeconomic factors that usually accompany dietary differences.

Studying this historical policy also offers useful contemporary guidance. The rationing system capped sugar intake at levels far below modern consumption norms, providing a rare population-wide test of intake thresholds that are otherwise difficult to evaluate causally. The results closely match the World Health Organization’s recommendation that added sugars remain below 10% of daily energy (about 50 g/day in a 2,000-calorie diet) and its advice that reducing intake to under 5% (around 25 g/day) yields further benefits [26]. During rationing, adults consumed roughly 40 g/day –comfortably within this recommended range. This alignment underscores the value of the episode as a real-world test case for understanding the long-term health effects of meeting, or even exceeding, modern public-health guidelines.

Building on this framework, our analysis yields three main findings. First, we find that the reduction in adult cancer incidence depends strongly on the duration of early-life exposure to sugar rationing. Hazard ratios fall progressively as exposure length increases, forming a clear dose–response gradient. For these fully exposed cohorts, cancer risk is markedly lower: liver (HR = 0.31; 95% CI: 0.18–0.49), rectum (HR = 0.60; 0.51–0.69), and prostate cancer (HR = 0.48; 0.43–0.55), alongside moderate but significant reductions for lung (HR = 0.59; 0.50–0.68) and breast cancer (HR = 0.64; 0.58–0.70). These patterns show that the protective effect strengthens with longer exposure to sugar rationing against cancer incidence.

Second, we identify a behavioural reinforcement channel linking early-life nutrition to long-term dietary habits. Using UK Biobank’s Oxford WebQ 24-hour dietary data [27], we find that those exposed to sugar rationing consumed less sugar, ate smaller food quantities, and had healthier, more diverse diets than cohorts conceived just after rationing ended. These patterns indicate that early nutritional scarcity durably recalibrated taste formation. This interpretation aligns with evidence that early exposures shape flavour learning [28], that sugar activates reward pathways in addictive-like ways [2], and that early environments imprint lasting hedonic responses to sweetness [29].

Third, we explore the biological pathways through which early-life sugar exposure influences later-life health. Diabetes and hypertension are among the most common comorbidities accompanying cancer incidence and progression, reflecting shared metabolic and inflammatory mechanisms [30, 31]. To examine these connections, we re-estimate the effects of early-life sugar rationing on four major cardiometabolic diseases—hypertension, type 2 diabetes, obesity, and heart failure, following [32] and [25]. We find consistent declines in risk with longer exposure, including approximately 20% lower hypertension and cardiovascular disease and 30–35% lower type 2 diabetes risk among rationed cohorts. These patterns are consistent with reduced activation of insulin and insulin-like growth factor (IGF-1) signalling, chronic inflammation, and lipid dysregulation – key pathways linking excess sugar to both metabolic and cancer risks [5, 6, 33]. Complementing these findings, we present epigenetic evidence suggesting slower cellular ageing among rationed cohorts. Individuals exposed to rationing from conception through at least 19 months postnatally exhibit 0.05 standard deviations longer leukocyte telomere length (LTL) (*p <* 0.05), equivalent to about 2.2 fewer years of biological ageing [34–36]. Since telomere attrition reflects cumulative oxidative and inflammatory stress, this molecular pattern provides a plausible biological link between early-life sugar restriction, attenuated metabolic strain, and reduced long-term disease susceptibility. Together, these findings indicate that early-life sugar exposure influenced adult health through both biological and behavioural pathways – mitigating metabolic stress, slowing cellular ageing, and shaping long-term consumption preferences.

Our study contributes to the existing literature in two important ways. First, we focus on cancer incidence as the primary health outcome, extending the same historical design of [25], [24] and [32], who study cardiometabolic and cardiovascular domains, to provide the first causal evidence linking early-life sugar exposure to cancer risk. Second, while prior studies interpret their results within the “fetal origins of disease” hypothesis, emphasizing biological programming during the first 1,000 days, we highlight a complementary mechanism that operates through the shift of long-term food preference. Specifically, we show that early-life sugar rationing induced a persistent shift in dietary preferences, reducing lifelong sugar consumption and influencing chronic disease risk through altered diet composition rather than physiological programming alone. This behavioural pathway is our key contribution: it demonstrates that early nutritional environments can shift hedonic preference and sustain health benefits far beyond early childhood.

## 2 Policy Background

The United Kingdom’s post-war rationing system provides a rare exogenous shift in early-life nutrition. From 1942 to 1953, sugar and sweets were strictly rationed under a centrally managed program designed to ensure equitable calorie distribution during scarcity. Each individual, including pregnant women and children aged two and above, was allotted about 8 oz of sugar per week (roughly 32 g per day) and 12 oz of sweets per month (roughly 11 g per day), redeemable via ration books at registered retailers. Children under two were not specifically allotted sugar or sweets, reflecting infant feeding practices that emphasized breastfeeding, formula, and age-appropriate foods such as grains, vegetables, and fruits. Because allocations were imposed by government decree rather than household choice, variation in sugar consumption was externally determined rather than driven by family preferences or socioeconomic status.

We document the changes of food consumption during the rationing period using the National Food Survey (NFS), which began in 1940 and tracked weekly food consumption for over 10,000 British households [37]. Rather than using the original survey records directly, we draw on the digitised and harmonised version of the NFS data compiled by [25], which provides quarterly, average per-person estimates of consumption across major food groups, including sugar, fats, meats, fruits, vegetables, and vitamins, between 1950 and 1960.

A natural concern is whether this shift reflects broader dietary or market-wide changes rather than sugar availability alone. Prior studies, [25] and [32], conduct several robustness checks, many of which we replicate here, to rule out common confounders.

First, although other food groups were derationed around a similar period, Appendix Figure S3 (replicating Figure S3 in [25]) shows no comparable rebounds in fats, dairy, cereals, fruits, vegetables, or meat; the 1953 jump is unique to sugar.

Second, total caloric intake rose only modestly—by about 100–150 kcal/day (<5% relative to 1950–1953 levels). Replications of Figure S1(c) in [25] and Supplementary Figure 2(c) in [32] show that sugar alone accounts for more than three-fourths of this increase, and sugar plus fat for nearly 90%, indicating that the overall calorie shift was driven primarily by changes in sugar availability.

Third, Appendix Figure S5 (replicating Figure S3 in [25]) shows stable sugar and general food prices, suggesting that government stockpiles prevented price pressures despite the surge in demand.

Fourth, rationing appears to have been enforced uniformly across socioeconomic groups. Supplementary Figure 2(d) in [32] shows similar pre–post changes in sugar intake by social class, consistent with limited access to black-market sugar and with the programme’s reputation for strong, universal enforcement.

Two additional observations follow from the descriptive evidence. First, the immediate post-1953 jump pushed adult sugar intake from roughly 40 g/day to around 80 g/day—well above modern World Health Organization guidance. The WHO recommends keeping added sugars below 10% of daily energy intake (about 50 g/day in a 2,000-calorie diet) and suggests further benefits from reducing intake to below 5% (about 25 g/day). Wartime rationing held intake at approximately 40 g/day, comfortably within these recommended thresholds. Second, the persistence of elevated sugar consumption after 1953, combined with stable patterns for other food groups, indicates a clear structural break rather than a temporary adjustment.

Taken together, these features, policy-driven variation, limited co-movement in other foods, stable prices, and uniform enforcement, make this historical episode a uniquely powerful setting. It offers a population-wide, quasi-experimental test of the long-term health effects of maintaining sugar intake within levels consistent with, or even stricter than, modern public-health recommendations.

## 3 Data and Methods

We use data from the UK Biobank, a large-scale prospective cohort study comprising over 500,000 participants aged 40–69 at recruitment (2006–2010) across 22 assessment centres in the United Kingdom. This dataset is particularly well suited for our study because its large sample size and age composition span cohorts conceived around the 1953 termination of sugar rationing, allowing us to identify individuals differentially exposed to the policy in early life. Moreover, the Biobank’s extensive linkage to genetic, health, and lifestyle data enables a comprehensive investigation of long-run biological and behavioural outcomes.

We exclude individuals who were: (1) born or living outside the UK at the time of the survey; (2) part of a multiple birth or adopted; (3) pregnant at the time of the survey; (4) those who withdrew their data from UK Biobank; and (5) those with missing covariates. Appendix Figure S2 presents the sample-selection flowchart.

Table 1 presents summary statistics for cohorts born between October 1951 and March 1956, comparing individuals conceived before the end of sugar rationing (born before June 1954) with those conceived afterward (born after June 1954). The two groups are broadly similar in demographic and socioeconomic characteristics, with only small differences in regional composition and smoking behaviour. Importantly, there are no statistically significant differences in parental cancer incidence between the rationed and derationed cohorts. This absence of systematic variation in family cancer history helps rule out concerns that genetic predisposition to cancer differs across exposure groups, reinforcing the validity of using the 1953 policy change as an exogenous early-life shock. Technical details on measurement protocols, quality control, and variable construction are provided in the Appendix S1.

**Table 1:** Characteristics of survey participants by exposure to rationing.

| Characteristic | Not rationed<br>*n* = 23, 782 | Rationed<br>*n* = 40, 979 | Difference | p-value |
| --- | --- | --- | --- | --- |
| Age at the latest survey | 55.9 | 57.9 | -2.0 | 0.000 |
| Male (%) | 43.5 | 43.8 | -0.3 | 0.469 |
| Place of birth |  |  |  |  |
| *England* (%) | 84.5 | 85.2 | -0.7 | 0.016 |
| *Wales* (%) | 4.8 | 5.0 | -0.2 | 0.356 |
| *Scotland* (%) | 9.7 | 8.9 | 0.8 | 0.001 |
| College (%) | 35.7 | 35.1 | 0.7 | 0.094 |
| Ever smoked (%) | 11.9 | 10.8 | 1.1 | 0.000 |
| BMI (mean) | 27.5 | 27.5 | -0.05 | 0.213 |
| Father's metabolic/chronic diseases and cancer incidence |  |  |  |  |
| Hypertension (%) | 25.2 | 22.5 | 2.7 | 0.000 |
| Diabetes (%) | 10.6 | 9.6 | 1.0 | 0.000 |
| Heart disease (%) | 33.1 | 33.6 | -0.5 | 0.210 |
| Prostate cancer (%) | 8.1 | 7.9 | 0.1 | 0.577 |
| Bowel cancer (%) | 5.4 | 5.9 | -0.5 | 0.017 |
| Lung cancer (%) | 8.8 | 9.1 | -0.4 | 0.126 |
| Mother's metabolic/chronic diseases and cancer incidence |  |  |  |  |
| Hypertension (%) | 35.5 | 34.1 | 1.4 | 0.001 |
| Diabetes (%) | 10.7 | 10.4 | 0.3 | 0.302 |
| Heart disease (%) | 20.9 | 21.9 | -0.9 | 0.005 |
| Breast cancer (%) | 8.2 | 8.1 | 0.2 | 0.544 |
| Bowel cancer (%) | 4.9 | 5.1 | -0.2 | 0.310 |
| Lung cancer (%) | 4.5 | 4.6 | -0.1 | 0.483 |

*Notes:* The table compares participants conceived before and after the sugar rationing cutoff. Characteristics are based on participants born between October 1951 and March 1956. Parental metabolic/chronic diseases and cancer incidence are self-reported indicators derived from responses to the questions “Was your biological mother/father ever diagnosed with stroke, heart disease, or hypertension?” and “What type of cancer did your father/mother have?” The rationed group comprises individuals conceived before the end of rationing (born before June 1954), while the non-rationed group comprises those conceived after rationing ended (born after June 1954). The final column reports p-values from tests of mean differences between the two groups.

## 4 Identification Strategy and Statistical analysis

Our empirical strategy follows the event-study framework of Gracner et al. [25], exploiting the abrupt end of sugar rationing in September 1953 as a natural experiment in early-life nutrition. The key comparison is between cohorts whose first 1,000 days of life, the critical developmental window, occurred before versus after this policy shift. Individuals conceived after September 1953 (born July 1954 or later) form the never-rationed group, having experienced a sugar-abundant environment throughout early development. Those conceived earlier (born before July 1954) provide the rationed comparison group, whose early-life diets remained constrained at levels close to modern dietary guidelines. Our analytic sample includes 64,761 adults (40,979 rationed; 23,782 non-rationed) born between October 1951 and March 1956. Because rationing ended sharply on a single date and applied uniformly nationwide, exposure is plausibly exogenous to family background and individual characteristics.

To capture variation in exposure intensity, we group individuals by the length of rationing they experienced: in utero only, in utero plus 6, 12, 18, or 24 months postnatally, or never exposed. The six-month intervals correspond to key stages in infant nutrition. Around six months of age, infants typically begin shifting from exclusive breastfeeding to supplementary and then predominant solid-food consumption, making this a natural breakpoint for how rationing could affect sugar intake. Figure 2 illustrates the rationing timeline and cohort distribution. These exposure indicators serve as the main explanatory variables, enabling comparisons between each exposure duration and the first fully never-rationed cohort—individuals conceived after September 1953 and born between July and December 1954. Treatment cohorts include those exposed in utero or early childhood for different lengths of time, while later-born post-rationing cohorts function as an additional check on broader secular trends.

**Figure 1:**
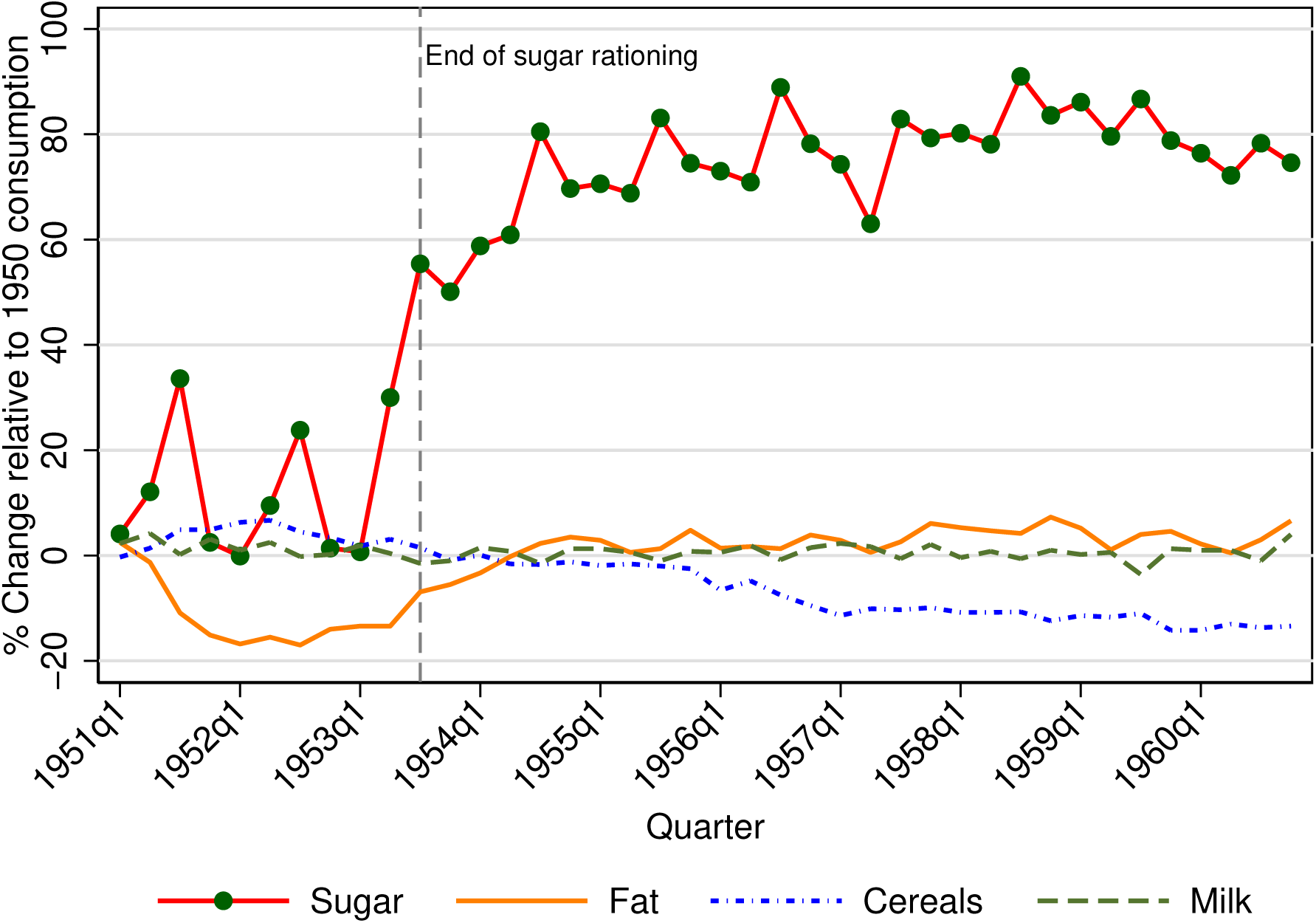
Trends in Food Consumption Patterns Around the End of Sugar Rationing *Note:* Based on NSF data. The figure shows quarterly percentage changes in sugar, cereal, milk, and fruit consumption relative to the 1950 average.

**Figure 2:**
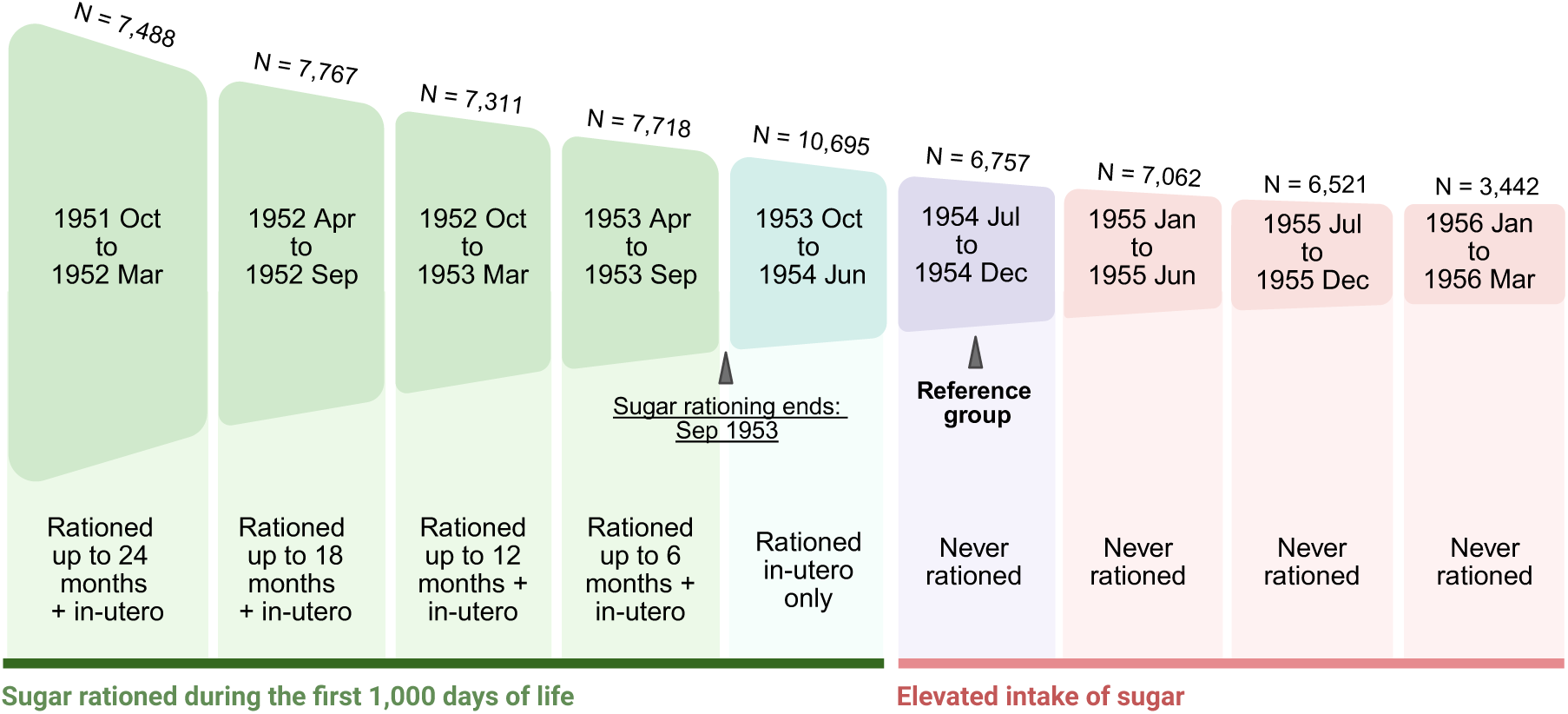
Timeline of survey participants’ exposure to rationing *Note:* Individuals exposed only in utero are shown in blue. Other rationed cohorts appear in green, and those never exposed are shown in red. The size of the shaded green bins reflects the duration of exposure. The earliest never-exposed cohort (born July–December 1954) serves as the comparison group and is used as the reference category in the regression analysis.

We include multiple never-rationed cohorts for two reasons. First, these cohorts provide a natural comparison group. Second, differences across the six-month post-policy cohorts allow us to test whether any observed patterns simply reflect secular trends or improvements in diagnostic practices that might make younger adults appear less healthy. Our identification assumption is that, conditional on covariates, outcomes among these non-rationed cohorts should exhibit no systematic trend.

To reduce compositional differences across birth groups, we adjust only for covariates that are exogenous before or at birth or that capture broad environmental context without lying on the causal pathway. These include age, sex, birthplace, birth-month seasonality, home-address latitude and longitude, the first ten genetic principal components, the Townsend deprivation index, and survey year. This set accounts for demographic, geographic, and genetic differences while avoiding adjustment for variables shaped by early-life nutrition. We therefore exclude early-life characteristics such as birth weight, breastfeeding duration, maternal smoking, and parental illness, which are potential mediators and are recalled 50–70 years later with substantial misclassification. For the same reason, we do not control for adult behaviours or conditions, smoking, alcohol use, BMI, diabetes, hypertension, or adult socioeconomic status, because these evolve over the life course and may themselves reflect early exposures. As a robustness check, we estimated extended models that add several family-background and individual factors, including parental illness, maternal smoking, participants’ own smoking status, educational attainment, and polygenic scores for relevant diseases. The results remain highly consistent across these specifications, indicating that our findings are not driven by omitted early-life family characteristics.

We estimate two sets of models to examine how early-life exposure to rationing affects biological ageing, dietary behaviour, and long-run health, with full expressions provided in Appendix S3. For continuous outcomes measured once in mid-adulthood, such as leukocyte telomere length, Granzyme B, healthy-eating index scores, and daily food weight, we use ordinary least squares. These estimates capture how early-life nutritional constraints manifest in biological ageing and dietary patterns by participants’ late 50s. Because individual sugar intake is unobserved, these regressions follow an intention-to-treat interpretation.

For clinical conditions, including cancer, diabetes, and cardiovascular disease, we estimate time-to-event models. The resulting hazard ratios capture how early-life exposure shifts the risk and timing of disease onset over the life cycle. We apply the same cohort structure as above, and post-rationing cohorts again serve as a validity check: conditional on controls, their hazard rates should remain flat relative to the reference group. We also compare survival curves for exposed and unexposed cohorts using a log-rank test.

As an additional validity test, we re-estimate the survival model using type 1 diabetes as a placebo outcome. Because this condition is largely driven by autoimmune and genetic factors, it should not respond to variation in early-life sugar access. Consistent with expectations, we find no cohort differences in incidence, reinforcing confidence in our identification strategy.

### Patient and public involvement

No direct patient or public involvement was undertaken for this study because no funding was available for patient and public involvement activities. The UK Biobank resource included extensive public consultation in its design. Although patients did not contribute to shaping the research question, outcomes, or study design, the project was initial inspired by informal conversations with a close family member who has relevant lived experience with the condition. After submission, we also asked a member of the public to read the manuscript and provide general feedback.

## 5 Main Results: Long-Term Health Outcomes

Our results demonstrate a strong, consistent, and dose-dependent protective effect of early-life sugar restriction on adult health, measured five to six decades after the nutritional shock.

### 5.1 Impact on Lifetime Cancer Incidence

We examine this possibility by estimating equation (2) using different types of cancer as outcomes, including liver, lung, rectum, breast (female only), and prostate (male only) cancers. Figure 3 presents the hazard ratios (HRs) relative the never-rationed reference group. For each cancer type, the line is flat before the policy shock, indicating no pre-existing differences, and then declines monotonically for cohorts exposed to rationing during their first 1,000 days. This clear dose-response pattern suggests that longer exposure to sugar restriction early in life led to progressively lower lifetime cancer risk. We further compare survival functions for exposed and unexposed cohorts using a log-rank test. Appendix Table S8 reports statistically significant differences for all cancer types, rejecting the hypothesis that both groups face the same cancer hazard.

**Figure 3:**
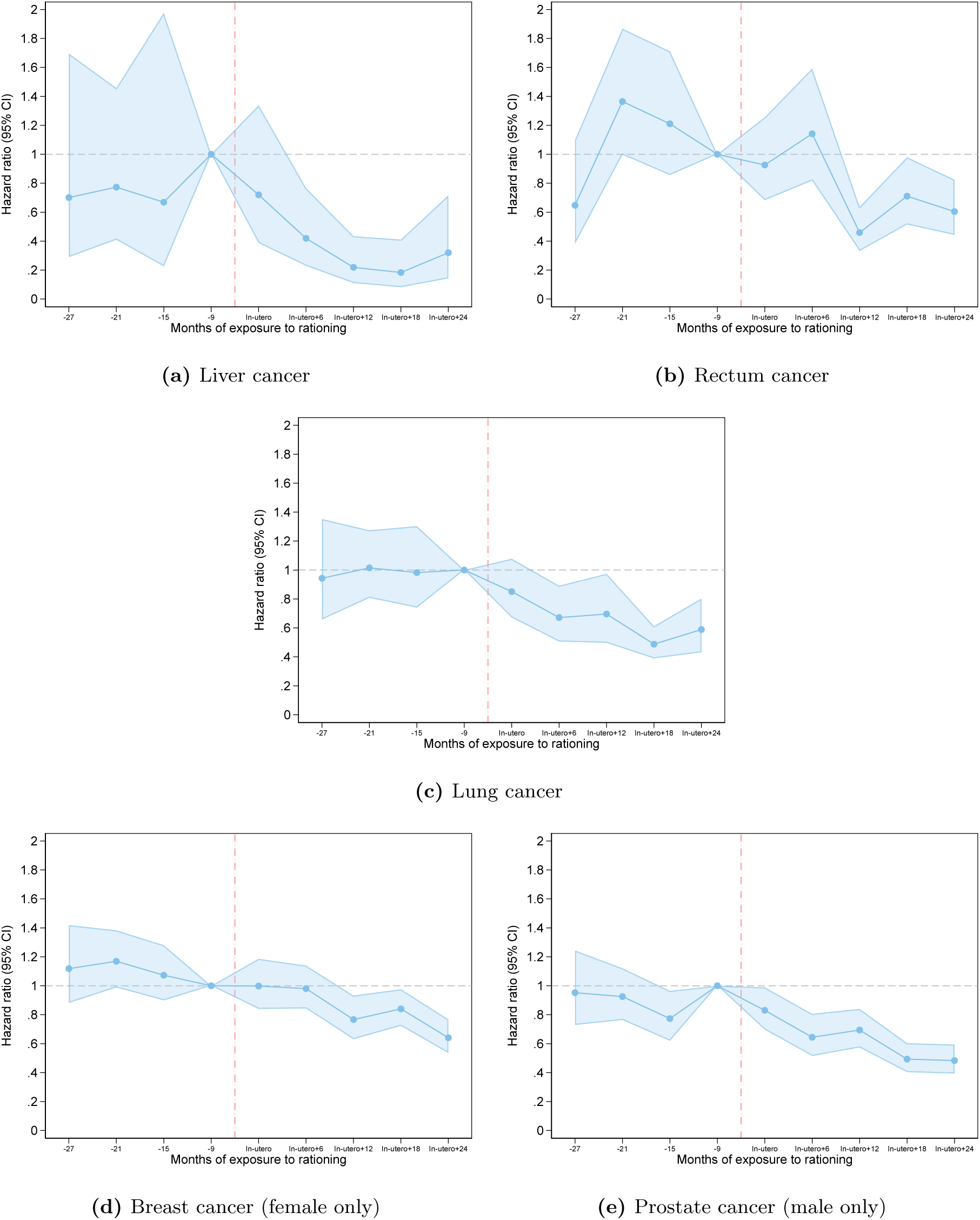
Association between early-life exposure to sugar rationing and cancer incidence in adulthood. Note: each panel reports hazard ratio (HR) estimates and 95% confidence intervals (shaded areas), relative to adults never exposed to rationing (born July–December 1954). HRs are estimated from equation (2), controlling for age, education, sex, birth location, calendar month of birth, and survey year. Groupings in this figure are consistent with those in figure 2. The x-axis measures exposure duration, with negative values denoting months since rationing ended at birth. The vertical dashed line separates the rationed group (conceived before the end of rationing; born before June 1954) from the derationed group (conceived after rationing ended; born after June 1954).

#### Digestive Cancers (Liver and Rectum)

The strongest and most precise effects appear for digestive cancers. Figure 3a shows liver cancer risk declines by nearly 70 %, consistent with biological and epidemiological evidence linking high fructose intake to hepatic insulin resistance, de novo lipogenesis, and non-alcoholic fatty liver disease—key precursors of hepatocellular carcinoma. Excess dietary sugar also elevates oxidative stress and promotes hepatic inflammation, accelerating carcinogenic pathways [14, 38]. The results therefore suggest that early-life exposure to a low-sugar environment prevented metabolic programming that would otherwise predispose individuals to liver cancer.

Figure 3b shows that rectum cancer exhibits a similarly strong dose–response pattern. Cohorts conceived before rationing (e.g., *−*21 or *−*15 months) show HRs statistically indistinguishable from 1.0, confirming the absence of pre-trends. Among those exposed during the critical 1,000-day window, risk falls sharply with longer exposure, reaching an HR of 0.60 (95 % CI: 0.42–0.85) for the “in - utero + 24 months” cohort. This finding aligns with evidence that high sugar diets disrupt the gut microbiome, promoting inflammatory phenotypes and reducing butyrate-producing bacteria that protect against tumour formation [39].^2^ An early-life environment that established a less inflammatory microbiome and healthier epithelial maintenance could thus confer lifelong protection against digestive cancers.

#### Respiratory Cancers (Lung)

Figure 3c shows that lung cancer exhibits a similar, though less pronounced, decline in risk. The longest-exposed cohort has a hazard ratio (HR) of 0.59 (95 % CI: 0.38–0.91), corresponding to a 41 % reduction in lifetime risk. Although the relationship between diet and lung cancer is indirect, systemic inflammation offers a plausible biological pathway. Chronic, low-grade inflammation is a well-established driver of carcinogenesis, even among non-smokers [40]. High sugar intake promotes such inflammatory states and contributes to insulin resistance, both of which increase oxidative stress and impair immune regulation. Because our models explicitly control for smoking status (never, former, current), the persistence of this association after adjustment suggests an independent metabolic or immune mechanism — potentially programmed during early development.

#### Hormone-Sensitive Cancers (Prostate and Breast)

The protective effect is especially pronounced for sex-specific, hormone-sensitive cancers. Figure 3d shows that the longest-exposed male cohort has a hazard ratio (HR) of 0.48 (95% CI: 0.31–0.74), while Figure 3e shows the corresponding female cohort with an HR of 0.64 (95% CI: 0.47–0.88). These findings are consistent with the insulin/IGF-1 hypothesis [5, 30], which posits that chronic exposure to high sugar levels elevates insulin and bioavailable IGF-1, stimulating cellular proliferation and inhibiting apoptosis in hormone-responsive tissues. Individuals conceived after the end of rationing, who were exposed to higher sugar intake starting in utero, may therefore have experienced sustained hyperinsulinemia and elevated IGF-1 throughout life, heightening their risk of hormone-dependent malignancies. Meta-analytic evidence supports this interpretation: existing studies document robust positive associations between sugar-sweetened beverage consumption and breast cancer as well as prostate cancer [41].

The monotonic decline in cancer incidence with longer exposure to sugar rationing reflects the cumulative protection conferred across successive developmental stages of the first 1,000 days. During gestation, limited maternal sugar intake prevents fetal hyperglycaemia and excessive insulin–IGF-1 signalling, reducing metabolic programming toward adiposity and inflammation [42, 43]. In infancy, breastfeeding provides a naturally low-sugar environment that shields against metabolic overstimulation, while delayed exposure to sweetened formulas or commercial baby foods limits the formation of early taste preferences for sugar [44, 45]. As children begin complementary feeding, sustained restriction further prevents early establishment of high-calorie, high-sugar diets that reinforce insulin resistance and chronic inflammation [41, 46]. Together, these stage-specific mechanisms compound over time, so cohorts exposed to rationing from conception through early childhood experienced the greatest protection against metabolic dysregulation, translating into the strongest reductions in adult cancer risk.

#### Cardiometabolic disease replication

To assess whether our cancer findings reflect a broader metabolic mechanism, we replicate the cardiometabolic analyses of [25] and [32]. Because cancer and metabolic diseases share pathways involving insulin signalling and chronic inflammation, similar protective patterns would indicate that early-life sugar restriction improved systemic metabolic health rather than exerting a cancer-specific effect. Figure S7 (a)-(d) show clear dose-dependent reductions in type 2 diabetes, hypertension, obesity, and heart failure; for example, the in-utero + 24-month cohort has hazard ratios of 0.72 for T2D and 0.84 for hypertension. These consistent gradients point to durable metabolic benefits.

#### Placebo replication (Type 1 diabetes)

Following [25], we also estimate effects on type 1 diabetes, a condition driven by autoimmune and genetic factors rather than metabolic pathways. As expected, the estimates are flat and centred around 1.0 (Figure S7e), providing a clean placebo result that supports a metabolic, rather than autoimmune mechanism.

## 6 Investigating the Mechanisms

Having established a causal link between the early-life sugar shock and long-term cancer incidence, we next investigate how this effect persists for over five decades. We test two complementary hypotheses: a behavioural pathway and a biological pathway.

### 6.1 The Behavioural Pathway: The “Sweet Tooth” Preference

To probe the behavioural mechanisms behind our findings, we test whether early-life exposure to sugar rationing is associated with different dietary choices in adulthood. Because the UK Biobank collects detailed dietary information only once (when participants are in their late 50s), our analysis captures a single midlife snapshot rather than dietary trajectories over the life course. Even so, adult dietary patterns remain informative for assessing whether early nutritional conditions shaped not only physiological programming, as emphasized by the fetal-origins framework, but also enduring preferences and consumption habits.

Figure 4a shows that individuals conceived after rationing ended consumed substantially more food by weight, on average 50–100 grams more per day (equivalent to roughly 1.5–3 kilograms of additional food per month or about 100–200 extra kilocalories per day), than those exposed in utero or during early childhood. At the same time, their diets scored roughly 1–1.5 points lower on the Healthy Eating Index and 0.5–1 point lower on the Food Diversity Score (Figure 4b). These differences are statistically significant (*p−value <* 0.05) and point to a clear divergence in both diet quantity and quality: post-rationing cohorts not only ate more but also maintained less balanced and less varied diets.

**Figure 4:**
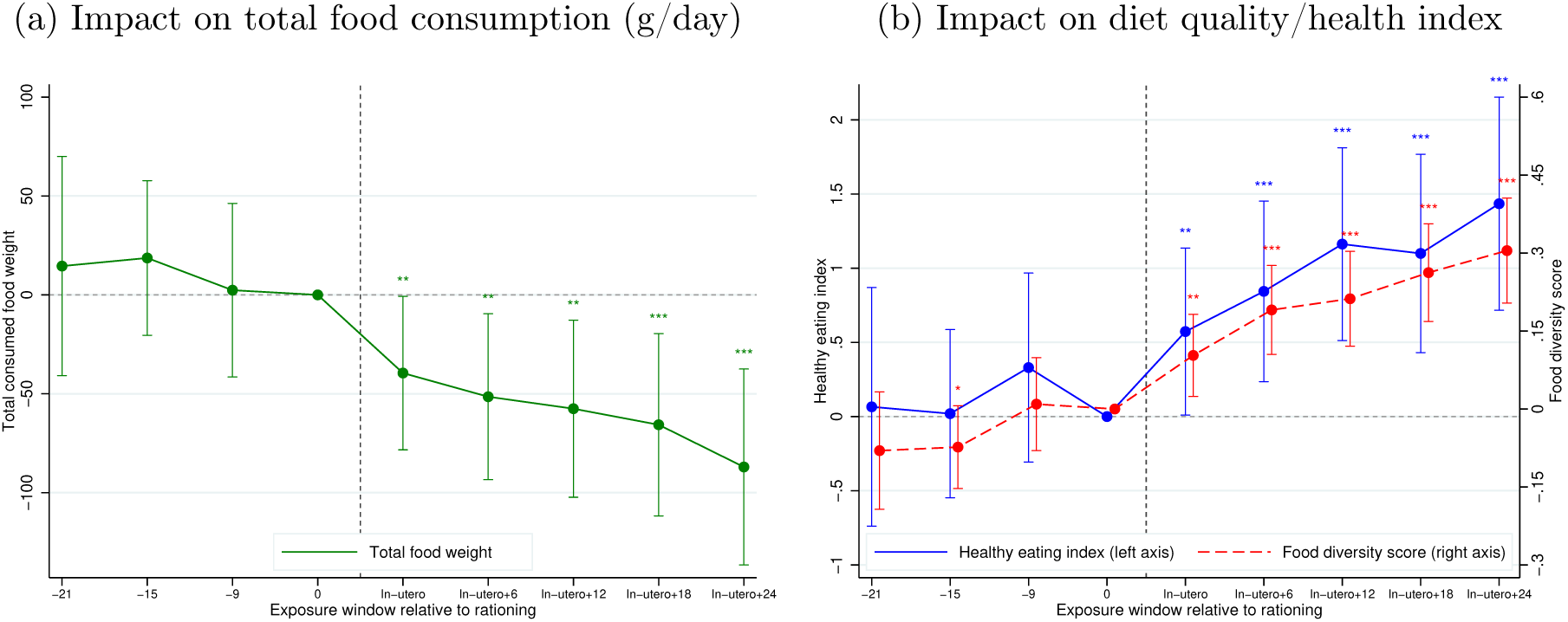
Impact of sugar rationing on diet patterns Note: Consumed food weight is based on self-reported intake of food, beverages, and water in the past 24 hours, providing a short-term proxy for longer-run diet quality and diversity. The Healthy Eating Index measures adherence to dietary guidelines across components (e.g., fruit, vegetable, protein, fat), with higher scores indicating healthier diets. The food diversity score reflects the number of distinct food groups con-sumed, indicating dietary variety and adequacy. Each point shows a cohort estimate comparing those exposed to rationing in utero, in utero plus 6/12/18/24 months, or not at all. The vertical dashed line separates the rationed group (conceived before the end of rationing; born before June 1954) from the derationed group (conceived after rationing ended; born after June 1954). /*/**/*** indicates significance at p smaller than 0.1/0.05/0.01/0.001.

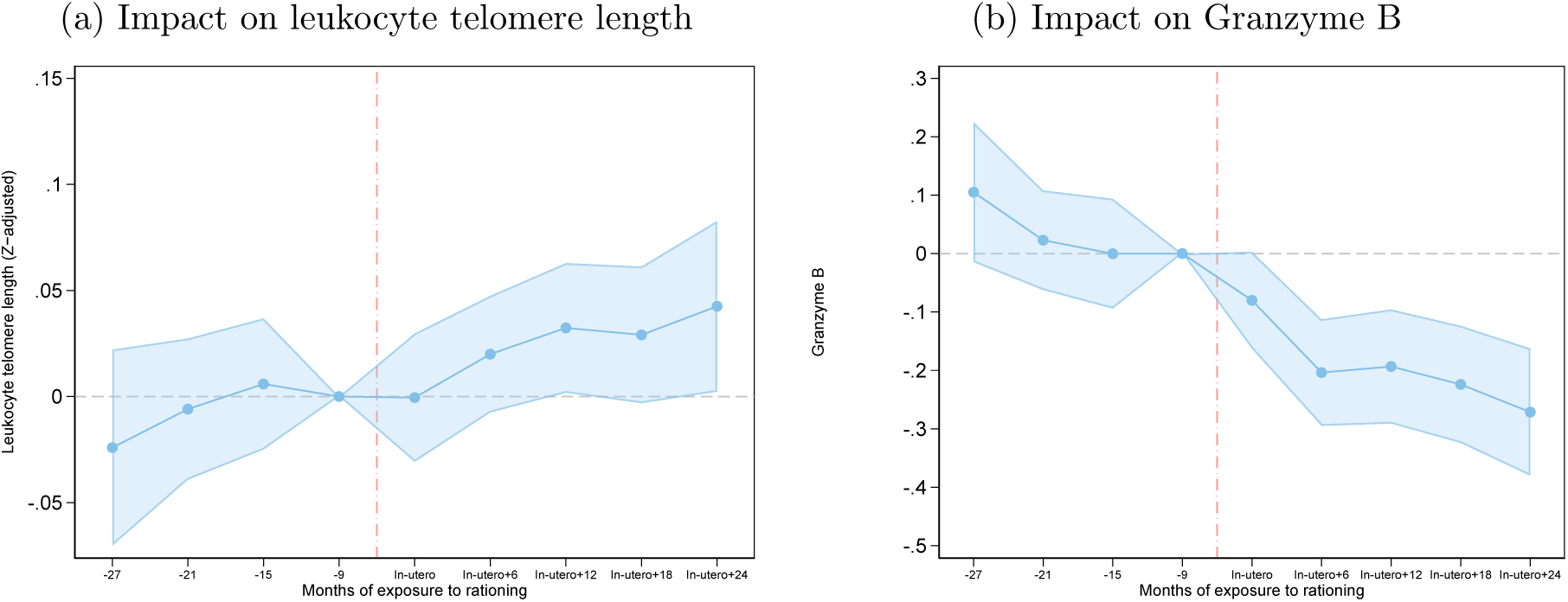
Note: Cohort groupings follow those shown in Figure 2. The x-axis represents the duration of exposure to sugar rationing, with negative values indicating the number of months elapsed since rationing ended at birth. Estimates are based on equation (1), controlling for age, education, sex, birth location, calendar month of birth, and survey year. The vertical dashed line separates the rationed group (conceived before the end of rationing; born before June 1954) from the derationed group (conceived after rationing ended; born after June 1954).

Disaggregating total intake by category (Figure S6) shows that cohort differences are driven mainly by sugar consumption. Rationed cohorts consumed substantially less sugar decades later, with smaller declines for fat and protein—consistent with the sugar–fat composition of confectionery and other ultra-processed foods. In contrast, starch and fibre intake remained stable, suggesting that rationing shaped not only energy intake but also long-term preferences for sweet, energy-dense foods.

The persistence of these differences supports a behavioural reinforcement mechanism: early nutritional experiences appear to calibrate taste and reward systems, influencing later dietary behaviour. Our findings indicate that the early-life environment generates a lasting “hedonic shift”—a change in the baseline of food reward sensitivity that biases individuals toward or away from highly palatable, sugar-rich foods. The first 1,000 days are a critical period for the formation of taste preferences and the maturation of dopaminergic reward pathways [28, 29]. During this window of neural plasticity, repeated exposure to high-sugar foods may “hijack” the brain’s reward system, centred in the ventral tegmental area and nucleus accumbens, by upregulating dopamine response to sweetness while dampening reward from less-sweet, nutrient-dense foods [2, 47, 48]. Once established, this hedonic calibration tends to persist, shaping food choices throughout life.

This behavioural mechanism adds an important dimension to the existing literature. Prior work by [25] and [32] focuses on physiological programming effects confined to the first 1,000 days—the so-called “fetal origins of disease” hypothesis—emphasizing metabolic and endocrine pathways. We complement and extend this framework by documenting a parallel behavioural channel: early nutritional environments also shape lifelong preferences through hedonic and sensory conditioning. Recognizing this mechanism broadens the scope for policy intervention. Interventions that moderate early exposure to hyper-palatable foods or incorporate structured taste education during infancy and early childhood could recalibrate food reward sensitivity, improving dietary quality and reducing chronic disease risk over the life course.

### 6.2 The Biological Pathways: Biological Imprinting and Cellular Ageing

We hypothesize that the nutritional shock left a direct and lasting biological imprint on the body by altering the fundamental pace of cellular ageing. We use leukocyte telomere length (LTL) as a well-established, integrative biomarker of biological ageing. Telomeres are protective nucleotide caps at chromosome ends that progressively shorten with each cell division. Although telomere shortening is a natural part of ageing, it is accelerated by metabolic stress, oxidative stress, and chronic inflammation. High-sugar diets are a major source of oxidative stress through the accumulation of advanced glycation end-products (AGEs) and mitochondrial overload, both of which directly damage telomeric DNA that is highly vulnerable to single-strand breaks [49]. Shorter telomeres are predictive of multiple age-related diseases, including cancer and cardiovascular disorders, and are strongly associated with higher all-cause mortality [50].

To capture the inflammatory pathway linking metabolism to cellular ageing, we examine circulating levels of Granzyme B, a cytotoxic enzyme secreted by T cells and natural killer (NK) cells to eliminate infected or malignant cells. While essential for immune defence, chronically elevated Granzyme B reflects a state of immunosenescence—an aged or exhausted immune system characterized by persistent, low-grade inflammation. Such chronic immune activation, driven by long-term metabolic stress from high sugar intake, accelerates T cell exhaustion and weakens immune surveillance—the process through which the immune system identifies and eliminates pre-cancerous cells [51–53].

Figure 5a reports event-study estimates of the effect of early-life sugar rationing on adult leukocyte telomere length (LTL). The dashed green line plots the estimated coefficients from equation 1, with 95% confidence intervals. Each estimate reflects the mean Z-adjusted LTL for a given birth cohort, relative to the reference group of never-rationed individuals born between July and December 1954. Estimates for cohorts conceived after the end of sugar ration show no systematic differences from the reference group, suggesting the absence of differential pre-trends. Cohorts exposed to rationing exclusively in utero exhibit no statistically significant effect on LTL. By contrast, cohorts with longer exposure durations display progressively larger increases in LTL. The largest increase is observed among individuals exposed from conception through at least 19 months postnatally, who exhibit 0.05 standard deviations longer LTL (*p <* 0.05). Based on UK Biobank estimates that LTL shortens by approximately 0.023 SD per year of chronological age, this effect corresponds to about 2.2 fewer years of age-related attrition [34]. This mapping is consistent with independent longitudinal studies reporting 15–30 base-pair annual shortening against an inter-individual standard deviation of 600–700 base pairs, reinforcing the interpretation of LTL as a biomarker of biological ageing [35, 36].

A near-symmetric pattern is observed for Granzyme B in Figure **??**: Granzyme B is a cytotoxic enzyme released by activated T cells and natural killer cells, and elevated circulating levels are considered a marker of chronic inflammation and immune ageing. Individuals conceived during the rationing period exhibit substantially lower levels in adulthood, consistent with a less inflamed, “younger,” and more resilient immune profile. Together, these results depict a coherent biological mechanism whereby early-life exposure to a low-sugar environment reduced metabolic stress, slowed cellular ageing, and preserved immune function into adulthood. In contrast, those from never-rationed cohorts experienced early metabolic overload that accelerated telomere attrition and promoted a pro-inflammatory, immunosenescent phenotype – compromising immune surveillance and increasing lifetime susceptibility to chronic disease.

## 7 Discussion and Conclusion

### 7.1 Summary and Synthesis of Findings

This study provides, to our knowledge, the first causal evidence linking an early-life shock in sugar availability to adult cancer risk and the fundamental pace of biological ageing. By exploiting the 1953 termination of UK sugar rationing as a natural experiment, we isolate exogenous variation in early-life exposure to sugar rationing, thereby overcoming the confounding factors that have long constrained observational research.

The findings reveal a coherent and multidimensional picture. Early-life exposure to sugar restriction causally reduced the adult incidence of multiple major cancers, including rectum, prostate, breast, and lung cancers, as well as a range of cardiometabolic diseases such as type 2 diabetes, hypertension, obesity, and heart failure. Beyond these disease outcomes, we uncover a lasting behavioural mechanism: early-life nutritional restraint appears to recalibrate taste preferences, a process of hedonic programming that leads to a persistently healthier, lower-sugar diet even five decades later. Parallel to this behavioural adaptation, we also identify a complementary biological pathway shaped by early-life nutritional programming. The early-life sugar shock appears to have permanently slowed the pace of cellular and immune ageing, as evidenced by significantly longer telomeres and a lower inflammatory profile in adulthood. The estimated difference of roughly 0.05 standard deviations in leukocyte telomere length corresponds to more than two fewer years of biological ageing.

Excessive sugar consumption can accelerate both disease progression and biological ageing through interconnected metabolic and inflammatory pathways (Figure 6). High sugar intake increases insulin and IGF-1 signalling, which promotes cell proliferation and suppresses apoptosis, thereby heightening cancer susceptibility. At the same time, sustained hyperinsulinemia and oxidative stress trigger chronic low-grade inflammation, marked by elevated circulating cytokines such as IL-6, TNF-*α*, and C-reactive protein. This pro-inflammatory state contributes to type 2 diabetes and hypertension and also speeds telomere attrition and cellular senescence, leading to faster biological ageing. Early-life sugar rationing, by preventing the activation of this metabolic–inflammatory cascade, likely produced a lasting physiological “reset” that protected multiple systems throughout adulthood.

**Figure 6:**
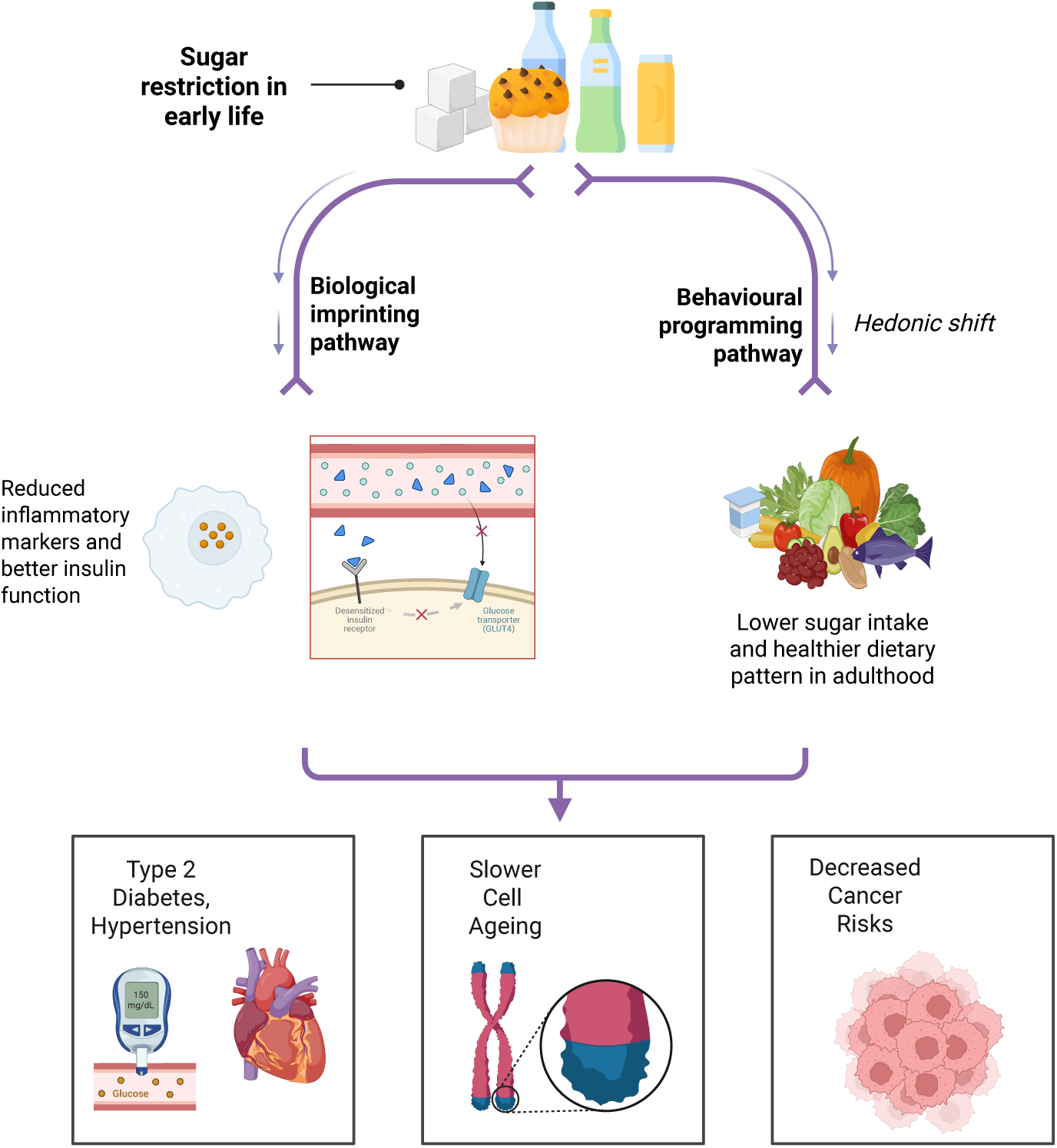
Biological pathways linking sugar intake to cancer risk, cardiometabolic disease, and cellular ageing.

These mechanisms are likely intertwined and mutually reinforcing. The initial biological imprint may have established a more favourable metabolic set point – characterized by higher insulin sensitivity, reduced oxidative stress, and lower inflammatory activation – while the behavioural pathway subsequently reinforced this advantage by reducing lifetime exposure to dietary sugar. The interaction of these processes yields the substantial, durable health differences we observe half a century later: a combination of improved metabolic regulation, reduced cumulative inflammatory burden, and slower biological ageing.

### 7.2 Limitations

Although this study benefits from a strong quasi-experimental design, a large population sample, and rich linked biological data, several limitations warrant discussion.

First, while the natural experiment provides plausibly exogenous variation, it is not a true randomized controlled trial. We cannot fully exclude unobserved differences between cohorts. Families who conceived immediately after the 1953 policy change may have differed systematically (e.g. in optimism, fertility planning, or post-war economic expectations) from those who conceived earlier. Nevertheless, the absence of any pre-trends in outcomes and the null effects on placebo conditions such as type 1 diabetes suggest that such unobserved factors are unlikely to explain our findings.

Second, the number of cancer cases is limited, particularly for certain site-specific cancers such as liver cancer (115 cases) and rectal cancer (374 cases). These relatively small sample sizes lead to imprecise point estimates and constrain our ability to examine potentially important heterogeneity by gender or ethnicity.

Third, the UK Biobank is known to suffer from a “healthy volunteer” bias, as participants are generally healthier and more educated than the broader UK population. However, this bias is unlikely to threaten our internal cohort comparisons. Unless selection probabilities varied discontinuously by six-month birth cohort in a manner correlated with the 1953 shock, the estimated differences across cohorts should remain unbiased.

Fourth, the 24-hour dietary recall used to measure adult diet provides only a cross-sectional snapshot and cannot capture lifetime consumption patterns. We therefore interpret it as a proxy for persistent dietary preferences rather than as a direct measure of lifetime intake. To the extent that this introduces classical measurement error, the bias would attenuate our estimates toward zero, suggesting that the true behavioural effect is likely larger than what we observe.

### 7.3 Policy Implications and Conclusion

In the United Kingdom, average free-sugar intake remains high—about 10.0% of total energy for adults and 10.5% for children, despite the recommended limit of 5% [54]. Globally, sugar consumption is projected to rise further, reaching roughly 22.5 kg per person per year by 2032 and about 23.1 kg by 2034 [55]. These figures highlight a persistent gap between current intake and public-health guidance across countries at all income levels.

The historical rationing period provides a powerful benchmark in this context. Wartime allocations kept adult sugar intake at roughly 40 g/day—a level that sat comfortably within the World Health Organization’s recommendation to keep free sugars below 10% of daily energy (about 50 g/day in a 2,000-calorie diet) and close to its more stringent 5% target (about 25 g/day). As consumption jumped to about 80 g/day immediately after rationing ended, intake moved well above these thresholds. This contrast offers a rare population-wide test of both compliance with, and departure from, modern nutritional guidelines.

The relevance is immediate. The first 1,000 days of life constitute a critical developmental window, yet today infants and young children are routinely exposed to diets that exceed recommended sugar levels. Our evidence indicates that early-life metabolic restraint, such as the low-sugar environment created inadvertently by wartime controls, can have durable benefits for cardiometabolic health, biological ageing, and cancer risk. In a world where sugar exposure is chronically high from infancy onward, the potential gains from reducing early-life intake are therefore substantial.

The policy implications follow. When baseline intake already exceeds optimal levels, early-life interventions have especially large payoff. Product-level regulation, such as mandatory limits on sugar content in infant formula, baby foods, and toddler snacks, would reduce exposure during the most sensitive developmental stage. Population-wide measures, including taxes on sugar-sweetened beverages, reformulation incentives, and front-of-pack warning labels, can shift the entire distribution of intake and indirectly protect the youngest cohorts.

## Author Contributions

CZ and WZ designed the study. CZ collected the data. CZ and WZ contributed to the analysis or interpretation of the data. WZ prepared the first draft of the manuscript. All authors contributed to the interpretation of the results and critical revision of the manuscript for important intellectual content and approved the final version of the manuscript. WZ is the guarantor. The corresponding author attests that all listed authors meet authorship criteria and that no others meeting the criteria have been omitted.

## Acknowledgment

The team thanks the volunteers, field staff, and data managers of the UK Biobank and the National Food Survey for their contributions. We are especially grateful to Tadeja Gracner and her co-authors for digitising the National Food Survey data and making it publicly accessible through their work.

## Data Availability statement

This research has been conducted using the UK Biobank Resource under application No. 89068. Data are available in a public, open access repository. Data from the UK Biobank are available to researchers on application.

## Transparency

The lead author (the manuscript’s guarantor) affirms that the manuscript is an honest, accurate, and transparent account of the study being reported; that no important aspects of the study have been omitted; and that any discrepancies from the study as planned (and, if relevant, registered) have been explained.

## Dissemination to participants and related patient and public communities

We plan to increase the impact of this research through several routes. First, UK Biobank will communicate key findings through its established participant outreach channels. Beyond this, we will share the results with nutrition specialists and oncologists at Addenbrooke’s Hospital, as well as local general practitioners, to help inform preventive advice related to cancer and cardiometabolic risk. We will also present the study at major academic conferences, integrate the findings into our teaching materials for students, and prepare a companion blog post explaining the results in accessible language. In addition, we will use informational social media platforms, such as to broaden public understanding. Finally, we will continue engaging with journalists and policy groups to highlight the implications of early-life sugar restriction for long-term health.

## Supplementary Appendix

### S1 Data Construction and Measurement

#### UK Biobank

The UK Biobank is a population-based cohort of 502,409 UK residents aged 40–69 at baseline, approved by the NHS National Research Ethics Service (Ref 11/NW/0382). Participants were recruited between March 2006 and July 2010 from 22 assessment centers across England, Scotland, and Wales. They provided socio-demographic, clinical, and lifestyle information, along with biological samples (blood, urine, saliva) and physical measurements, under written informed consent. This study uses data under application 89068.

#### Leukocyte telomere length (LTL

Our primary measure of biological ageing is LTL. Telomeres are DNA–protein complexes that cap chromosome ends, shorten with cell division, and are further eroded by inflammation, oxidative stress, and psychosocial stress [49, 56–59]. Shorter LTL is consistently associated with higher morbidity [60–62], mortality [63], and greater risks of Alzheimer’s disease and coronary artery disease in Mendelian randomization studies [64, 65]. In the UK Biobank, LTL was measured using quantitative polymerase chain reaction (qPCR), comparing telomere (T) to single-copy gene (S) copy number, yielding a T/S ratio proportional to mean telomere length. Valid LTL measures are available for 472,590 participants; we standardize the T/S ratio to mean 0, SD 1 [34].

#### Granzyme B

We use Granzyme B, an inflammatory activity marker that reflects both immunosenescence and chronic immune activation. Granzyme B is a cytotoxic serine protease secreted primarily by activated CD8^+^ T cells and natural killer (NK) cells to induce apoptosis in infected or malignant cells. Elevated circulating levels are indicative of heightened immune activation and systemic inflammation. Prior studies in older adults show that shorter leukocyte telomere length (LTL) is associated with higher Granzyme B concentrations, consistent with immune ageing and the senescence of cytotoxic lymphocytes [51–53]. In the UK Biobank, circu-lating Granzyme B was measured from stored EDTA plasma samples collected at baseline using a high-sensitivity Olink® Target 96 Inflammation panel, based on proximity extension assay (PEA) technology. Protein levels are expressed on a log_2_-scaled Normalized Protein eXpression (NPX) unit, enabling standardized comparison across individuals.

#### Dietary intake

Food consumption data come from the Oxford WebQ, a validated 24-hour web-based dietary recall introduced in 2009. The questionnaire covers 206 foods and 32 drinks, and UK Biobank nutrition experts convert reported intakes into nutrient values. Up to four recalls per person were collected for 210,977 participants. These surveys capture diet in late adulthood (typically age 50+), which we treat as a proxy for persistent dietary preferences. From these data, we construct two indicators of dietary quality: (1) a Healthy Eating Index (HEI), scored against dietary guidelines following [66, 67], in which higher scores reflect greater intake of fruits, vegetables, whole grains, fiber, and fish and lower intake of added sugars, saturated fat, sodium, and processed meat; and (2) a food diversification score, counting the number of major food groups consumed during the recall [68, 69]. Higher diversification reflects a more varied diet associated with better micronutrient adequacy.

#### Health outcomes

We examine a broad set of chronic disease outcomes spanning metabolic, cardiovascular, and cancer-related conditions, harmonized using ICD-10 codes (Supplementary Table S1). Diagnoses are drawn from linked NHS hospital admissions, cancer registries, primary care data, and mortality records, ensuring comprehensive longitudinal coverage of first-time disease events. We focus on key metabolic disorders, type 2 diabetes mellitus (E11), hyper-tension (I10), and obesity (E66), which capture major pathways connecting biological ageing to cardiometabolic function. Heart failure (I50) is included as a downstream cardiovascular endpoint. To assess cancer risks, we consider breast (C50), prostate (C61), lung (C33–C34), rectum (C20), and liver cancer (C22), with breast and prostate cancers treated as sex-specific. Follow-up extends to 2021–2022 for most diseases and to March 2023 for cancer and mortality registries.

We track two key metabolic outcomes, type 2 diabetes mellitus (E11) and hypertension (I10),which together capture major pathways linking biological ageing with cardiometabolic dysfunction. We also include obesity (E66) as a related metabolic disorder, and heart failure (I50) as a downstream cardiovascular endpoint.

To capture cancer risks, we consider breast (C50), prostate (C61), lung (C33–C34), rectum (C20), and liver cancer (C22). Breast and prostate cancers are sex-specific, while the others are common to both sexes. These outcomes allow us to assess whether slower biological ageing corresponds to lower incidence of both metabolic and oncologic diseases, reflecting the systemic influence of cellular maintenance processes on long-term health trajectories.

#### Control variables

In our main specification, we adjust only for covariates that are fixed before or at birth or that capture broad environmental context without lying on the causal pathway. These include age, sex, birth location, calendar month of birth, home-address lat-itude and longitude, educational attainment, the first ten genetic principal components, the Townsend deprivation index, and survey year. This set accounts for demographic, geographic, and genetic differences that may shape baseline health risks while avoiding adjustment for vari-ables influenced by early-life nutrition. We intentionally exclude early-life characteristics such as birth weight, breastfeeding duration, maternal smoking during pregnancy, and parental illness because these factors are potential mediators of the effect of sugar rationing and are measured using retrospective recall 50–70 years later, making them both conceptually downstream and highly prone to misclassification. We also do not control for adult behaviours and conditions, such as smoking, alcohol use, BMI, diabetes, hypertension, and adult socioeconomic status, be-cause these outcomes are shaped over the life course and may themselves reflect early-life sugar exposure. Adjusting for them would therefore create over-adjustment bias and attenuate the estimated treatment effects.

### S2 Sugar intake and price during the period

Figure S1 presents the timeline of food rationing between 1939 and 1954. Restrictions began to be lifted gradually after World War II, starting with bread and flour in July 1948, followed by biscuits, jam, and canned or dried fruits in 1950. While fats, rice, cheese, meat, milk, and cereals remained rationed until July 1954, sugar and sweets were de-rationed earlier, in September 1953. Their removal triggered an immediate, nutrient-specific shock: sales of sweets and chocolate rose by more than 150% within a year [23], and average adult sugar intake doubled—from about 41 g per day in early 1953 to nearly 80 g by late 1954. Among children, consumption of sugar and sweets more than doubled from pre-rationing levels, accompanied by a sharp deterioration in oral health [70–74]. By contrast, the end of rationing for other foods produced only minor adjustments in consumption, underscoring sugar’s unique role as the key driver of dietary change. All food rationing ended by July 1954.

### S3 Statistical Analysis

Our analysis is based on in two specifications.

#### Specification 1: Baseline Regression

We begin by estimating the average effect of the nutritional shock on later-life outcomes. This provides a benchmark for assessing whether expo-sure to rationing in early life has persistent biological consequences. Our baseline specification is an event-study design:

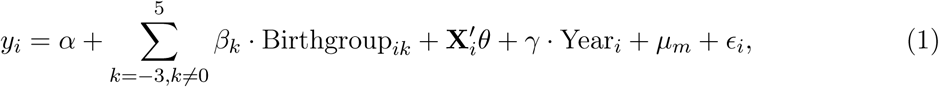

where the dependent variable *y_i_* measures long-run outcomes for individual *i*. Our primary outcomes are biomarkers of cellular ageing, most notably leukocyte telomere length (LTL). In addition, we examine dietary outcomes related to sugar, fat, and protein consumption, which we treat as potential mediating channels linking early-life nutrition to biological ageing.

The central explanatory variables are the cohort indicators Birthgroup*_ik_*, which capture differential exposure to sugar rationing by birth timing. The coefficients *β_k_* represent the av-erage effect of being in cohort *k* relative to the omitted reference group (*k* = 0), consisting of individuals born in 1954 Q3–Q4. Cohorts include treatment groups exposed to rationing for varying durations (*k* = 1*, . . .,* 5) and post-treatment cohorts conceived after rationing ended (*k* = *−*1*, −*2*, −*3).

Binary indicators for post-rationing cohorts help ensure comparability across birth groups. These later-conceived cohorts function as a natural control group and allow us to test for con-founding factors such as secular improvements in diagnostics or general time trends that could otherwise make younger cohorts appear biologically older. Our identification relies on the as-sumption that, conditional on covariates, outcomes for non-rationed cohorts should display no systematic trend. To mitigate compositional differences across birth groups, we control for sex, college attendance, birth-location coordinates, and the first five genetic principal components (**X***_i_*). We include calendar-month-of-birth indicators (reference: January) to absorb seasonal influences (*µ_m_*), and survey-year fixed effects to capture nationwide trends in outcome measure-ment and health reporting (Year*_i_*).

This baseline should be interpreted as an Intention-to-Treat (ITT) design: while individual confectionery consumption is unobserved, we estimate the impact of the policy shock itself. Archival evidence confirms a sharp increase in demand for sweets after derationing, although such purchases were excluded from the National Food Survey.

#### Specification 2: Health Outcomes

If the nutritional shock accelerates cellular ageing, the key question is whether this translates into higher incidence of chronic disease. To test this, we estimate a stratified proportional hazards model:

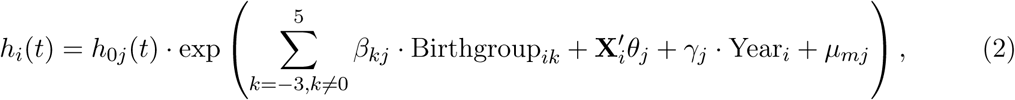

where *h_i_*(*t*) is the hazard of disease onset at time *t*, and *h*_0_*_j_*(*t*) the baseline hazard, modeled with a Gompertz distribution. As in the event-study, the coefficients *β_kj_* capture relative risks by cohort, benchmarked against the first never-rationed cohort (conceived after September 1953 or born July–December 1954). Our identifying assumption is that, conditional on controls, hazard rates among post-rationing cohorts should remain flat, keeping their hazard ratios near 1.

Together, these two specifications provide a comprehensive empirical strategy. Specification 1 establishes the average impact of early-life nutrition on biological ageing. Specification 2 further links these biological changes to long-run health, testing whether accelerated cellular ageing serves as a pathway from early-life shocks to adult chronic disease.

### S3.1 Impact on Cardiometabolic Disease

Given that cancer and cardiometabolic diseases share common etiological pathways, particularly insulin signalling, chronic inflammation, and obesity, we next examine whether the rationing shock exerted a similar protective effect on major metabolic outcomes. This analysis follows the empirical design of [25] and [32], who documented the long-term effects of sugar rationing on cardiometabolic health. However, our purpose differs from theirs. Whereas those studies fo-cused on cardiometabolic outcomes as endpoints of interest, we replicate their framework to test the biological mechanism underlying our cancer findings. Because cancer and cardiometabolic diseases are biologically intertwined through shared metabolic and inflammatory pathways, evi-dence of comparable protective effects across both would indicate that early-life sugar restriction improved systemic metabolic health rather than exerting a cancer-specific influence. Demon-strating such consistency provides crucial validation for the proposed biological channel.

The estimation results are reported in Figure S7, (a) - (d). Across all four outcomes, type 2 Diabetes (T2D), hypertension, obesity, and heart failure, we observe coherent, dose-dependent protective patterns. The In-utero + 24 m cohort exhibits a hazard ratio (HR) of 0.72 (95 % CI [0.58, 0.90]) for T2D and 0.84 (95 % CI [0.77, 0.92]) for hypertension, representing 28 % and 16 % reductions in lifetime risk, respectively, relative to the never-rationed cohort. Similar gradients are evident for obesity and heart failure, suggesting that prolonged exposure to early-life nutritional scarcity durably “programmed” higher baseline insulin sensitivity, healthier blood-pressure regulation, and overall metabolic resilience. The coherence of these results across the entire cluster of metabolic disorders demonstrates that early-life sugar restriction conferred broad, long-lasting, and systemic protection against cardiometabolic disease.

#### Placebo Tests

To rule out alternative explanations such as contemporaneous improve-ments in healthcare or post-war socioeconomic recovery, we performed placebo analyses in Fig-ure S7e. For type 1 diabetes (T1D), a disease driven by autoimmune and genetic factors rather than metabolic ones, the event-study estimates are flat, with all cohort HRs centred around 1.0 and no discernible trend. This null result aligns with expectations if the main effects arise specifically through metabolic channels.

---

**Figure S1:**
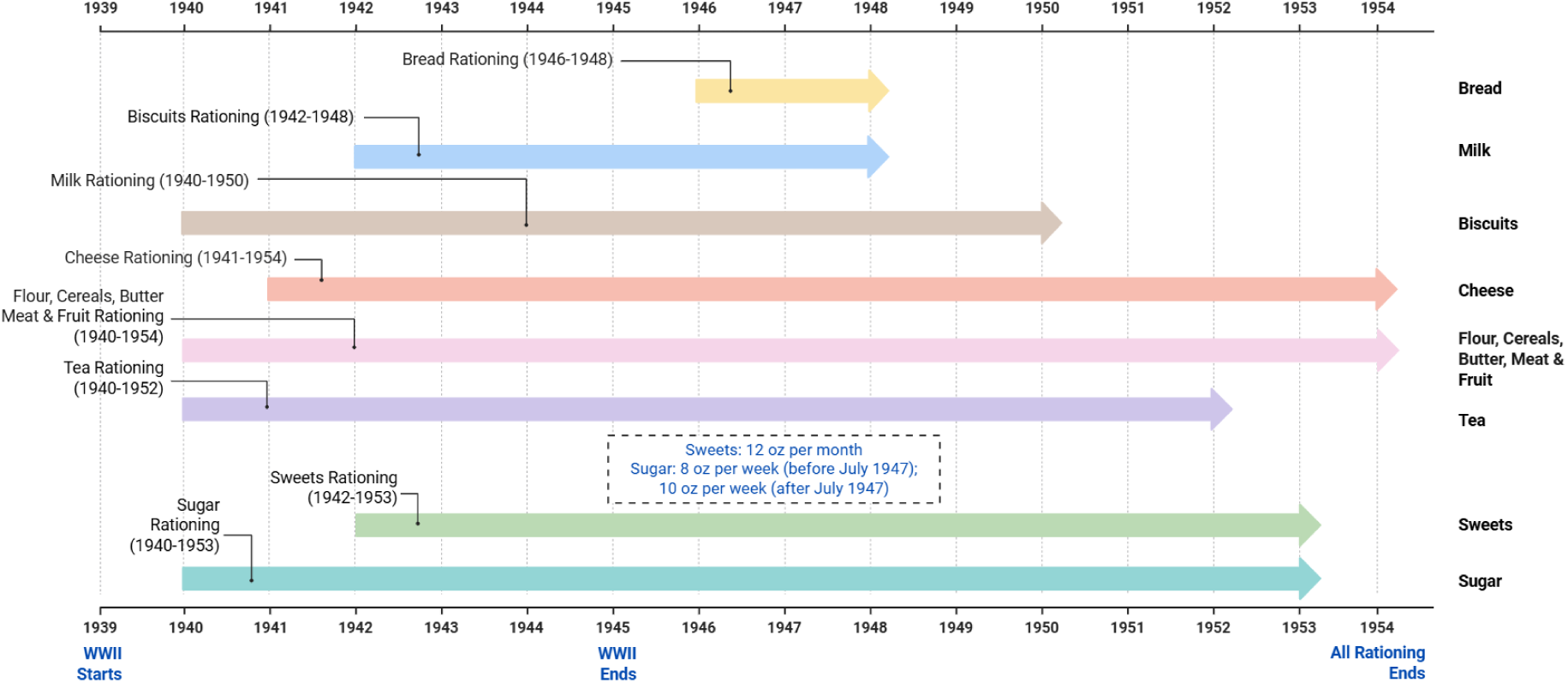
Food rationing timeline in the UK: 1939-1954

**Figure S2:**
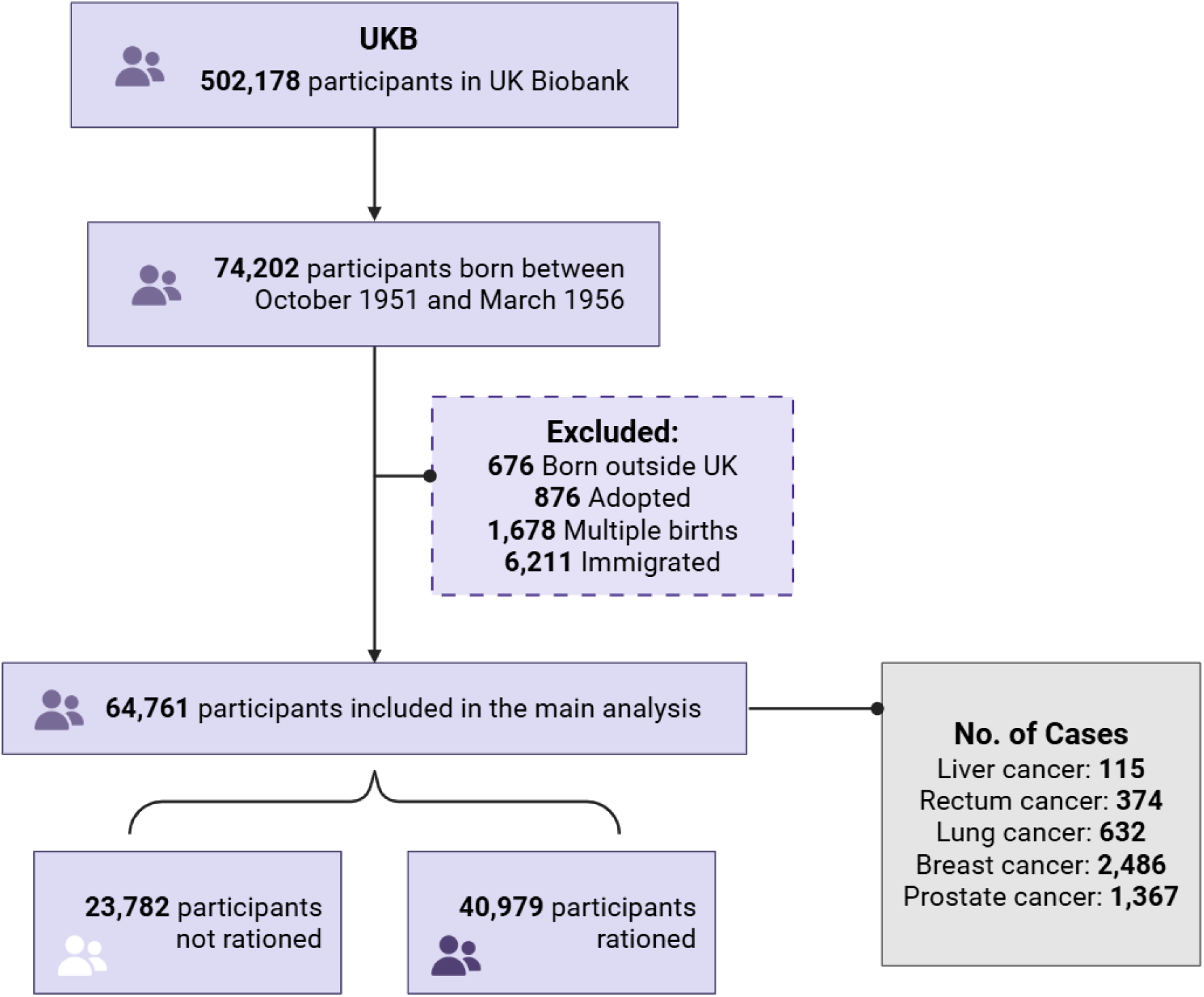
Flowchart of sample selection

**Figure S3:**
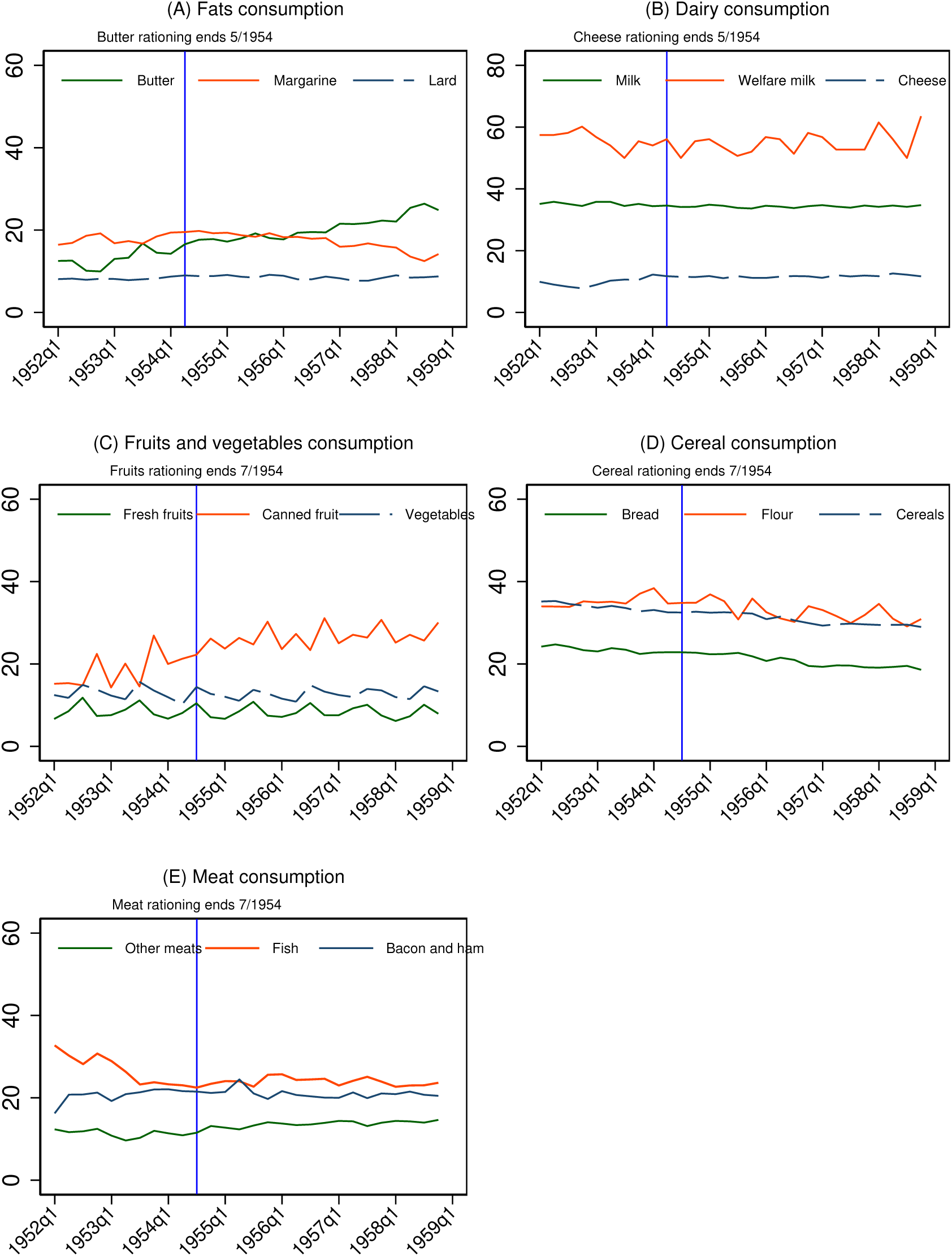
Quarterly consumption of fats, dairy, fruits and vegetables, cereal, and meat between 1952q1 and 1958q4 Note: this graph replicates Figure S3 in [25]. (A) Quarterly consumption of fats (butter, margarine, and lard). (B) Quarterly consumption of dairy (milk, cheese, milk on welfare/in schools). (C) Quarterly consumption of produce (fresh fruit, fresh vegetables, and canned vegetables and fruit). (D) Quarterly consumption of cereal (bread, flour, and cereals). (E) Quarterly consumption of meats (fish, bacon and ham, and other meats). Quarterly data on detailed food sub-categories is consistently available from 1952q1 through 1958q4. All quantities were originally reported in oz (or pints for milk) per head per week, which we converted into grams per day. Milk, fresh fruits, vegetables, bread and cereal are reported in 10 g grams per day to simplify visualization.

**Figure S4:**
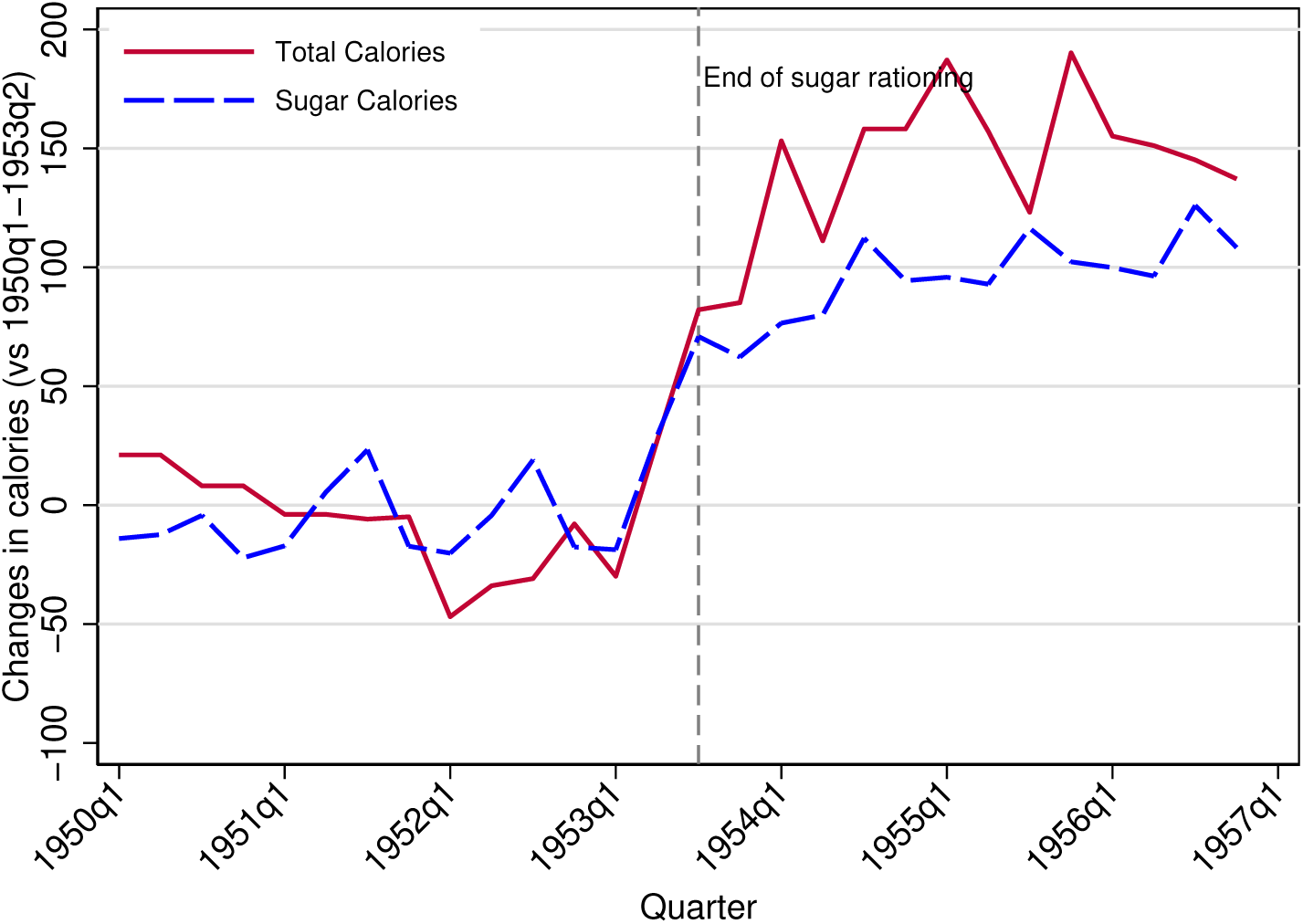
Calorie consumptions between 1950 to 1957 Note: this figure replicates Figure S1(c) in [25]. The blue line depicts total calories from all foods, while the red line shows calories from sugar alone.

**Figure S5:**
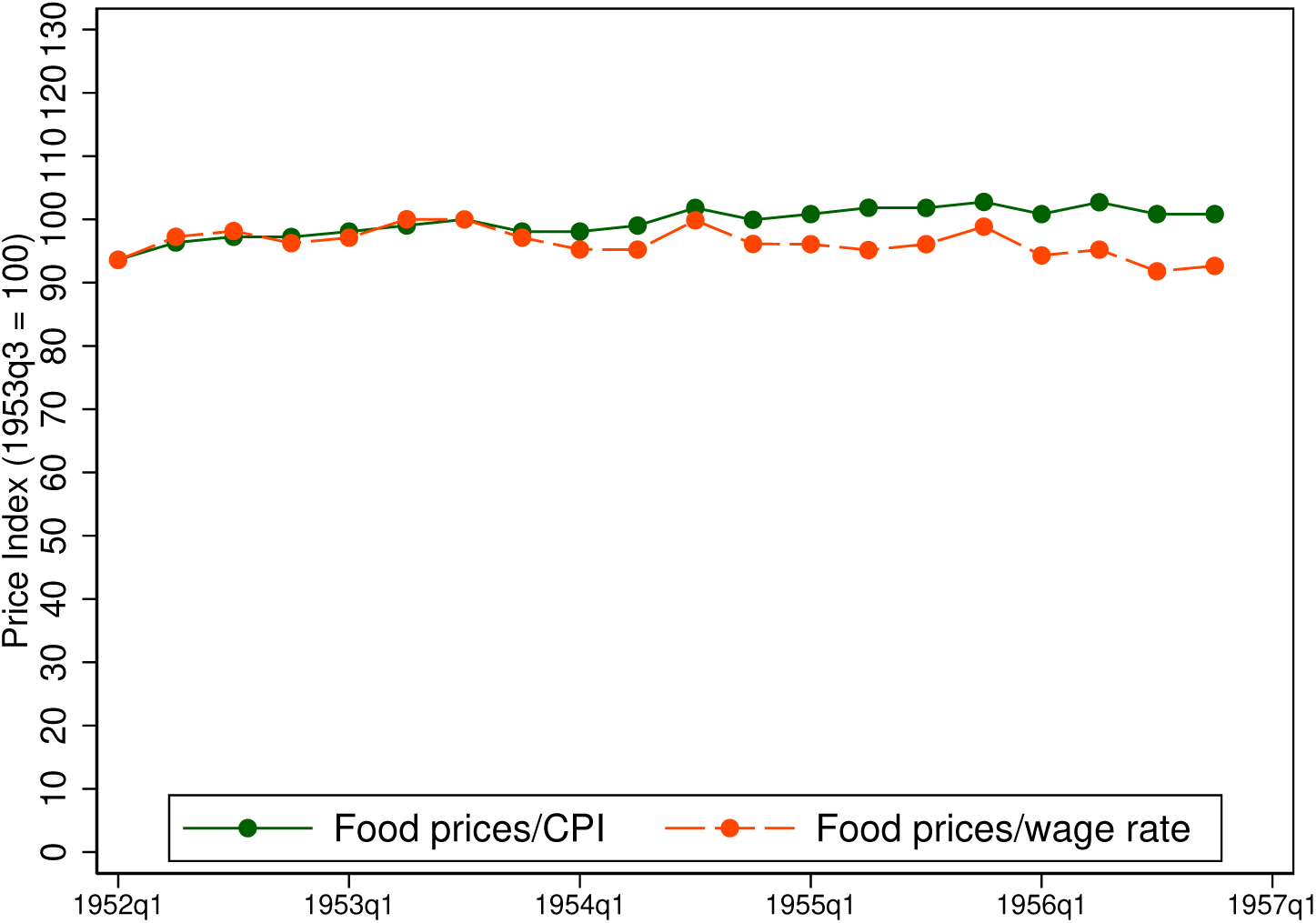
Food prices between 1952q1 and 1956q4 Note: this graph replicates Figure S4 in [25]. Food prices relative to general inflation and wage growth between 1952q1 and 1956q4 (1953q3=100). Quarterly data on prices were collected from historic NFS reports, where they were consistently available from 1952q1 through 1956q4.

**Figure S6:**
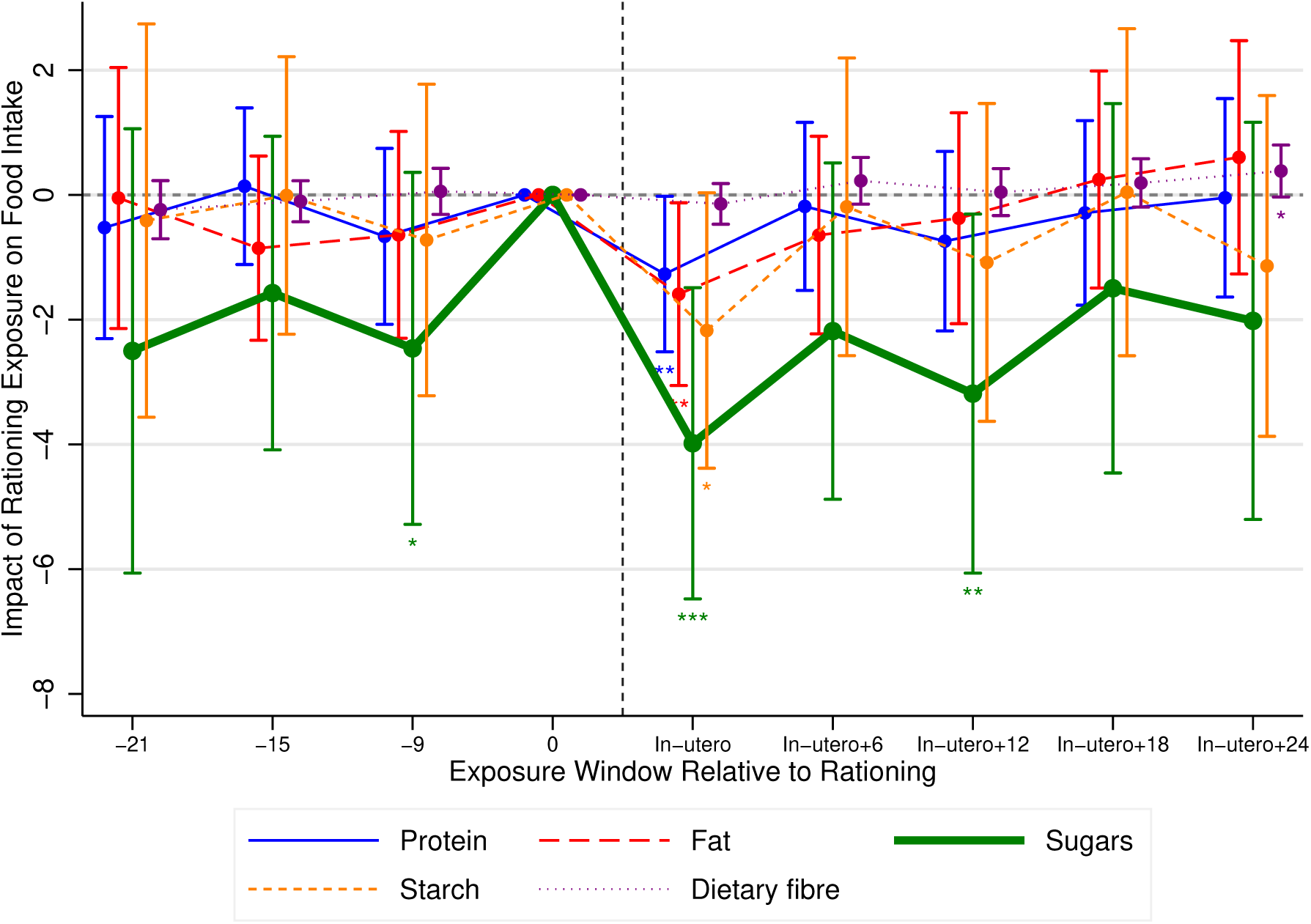
Impact of Rationing Exposure on Food Intake (by Category) Note: Each point shows a cohort estimate comparing those exposed to rationing in utero, in utero plus 6/12/18/24 months, or not at all. The vertical dashed line separates the rationed group (conceived before the end of rationing; born before June 1954) from the derationed group (conceived after rationing ended; born after June 1954). /*/**/*** indicates significance at p smaller than 0.1/0.05/0.01/0.001.

**Figure S7:**
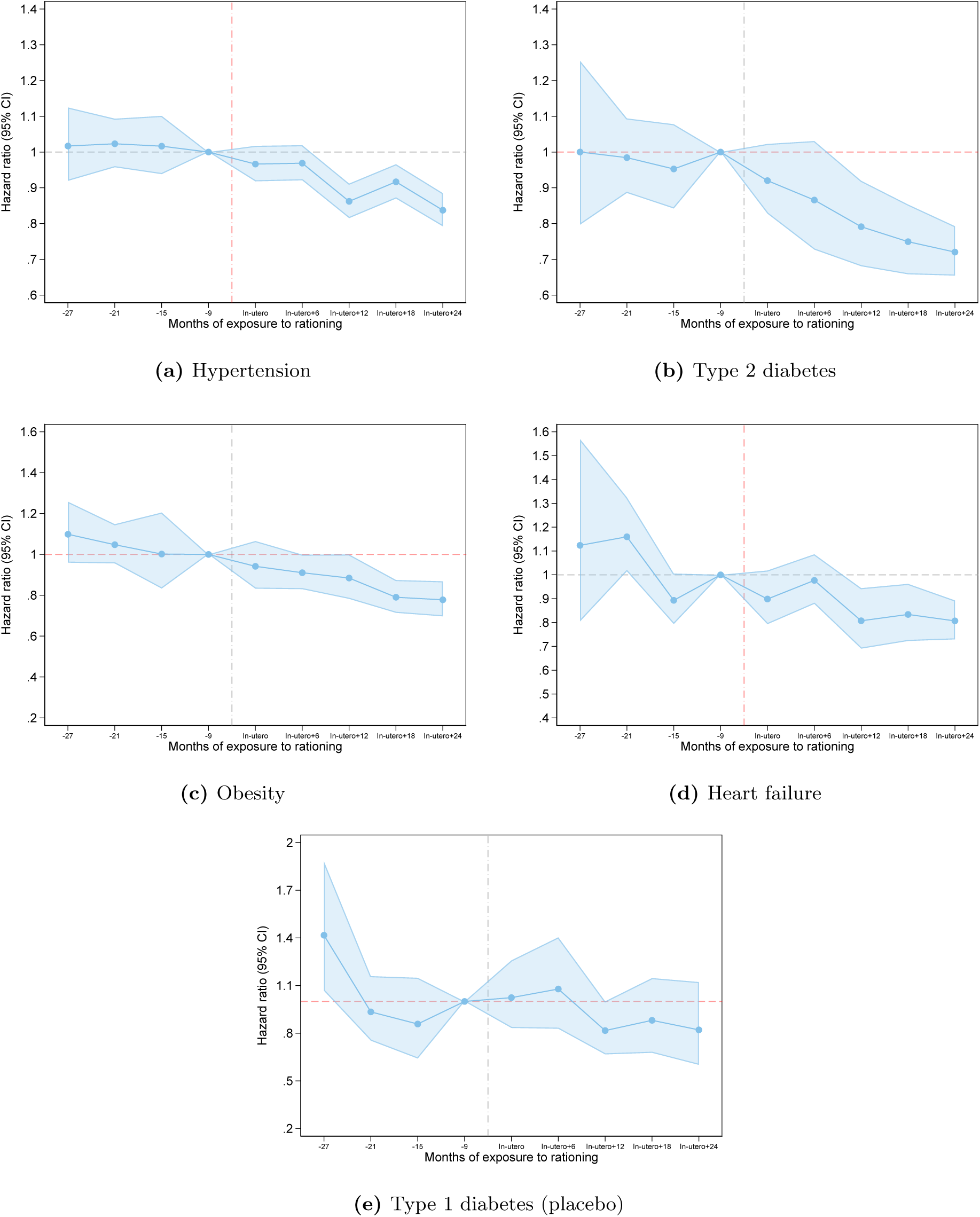
Replication of cardiometabolic outcomes as validation for shared metabolic pathways Note: this figure replicates the analysis of [32] and [25] for five cardiometabolic outcomes: hypertension, type 2 diabetes, obesity, heart failure and type 1 diabetes. Each panel shows hazard ratios (HRs) with 95% confidence intervals (shaded areas), relative to adults unexposed to sugar rationing (born July–December 1954). HRs are estimated from equation (2), controlling for age, education, sex, birth location, calendar month of birth, and survey year. The x-axis measures exposure duration, with negative values denoting months since rationing ended at birth. The red dashed line separates the rationed group (conceived before the end of rationing; born before June 1954) from the derationed group (conceived after rationing ended; born after June 1954).

**Table S1:** Disease outcome definitions.

| Disease outcome | ICD-10 codes | Number of cases | Sex specific |
| --- | --- | --- | --- |
| Breast cancer | C50 | 2,486 | F |
| Prostate cancer | C61 | 1,367 | M |
| Lung cancer | C33–C34 | 632 |  |
| Rectum cancer | C20 | 374 |  |
| Liver cancer | C22 | 115 |  |
| Type 2 Diabetes mellitus | E11 | 4,262 |  |
| Hypertension | I10 | 21,275 |  |
| Obesity | E66 | 2,171 |  |
| Heart failure | I50 | 1,548 |  |
| Type 1 Diabetes mellitus | E10 | 633 |  |

*Notes:* This table reports ICD-10 code definitions and the number of diagnosed cases for each disease outcome in the study sample. “Sex specific” indicates outcomes limited to either female (F) or male (M) participants.

**Table S2:** Hazard ratios (95% CI) for cancer incidence by exposure duration to sugar rationing.

| Exposure duration (months) | Lung<br>(1) | Breast<br>(2) | Liver<br>(3) | Prostate<br>(4) | Rectum<br>(5) |
| --- | --- | --- | --- | --- | --- |
| –27 months | 0.91<br>[0.65, 1.29] | 0.70<br>[0.29, 1.70] | 0.65<br>[0.38, 1.10] | 1.12<br>[0.88, 1.42] | 0.95<br>[0.73, 1.24] |
| –21 months | 0.98<br>[0.79, 1.22] | 0.77<br>[0.41, 1.46] | 1.36*<br>[1.00, 1.87] | 1.17*<br>[0.99, 1.38] | 0.93<br>[0.77, 1.12] |
| –15 months | 0.95<br>[0.73, 1.24] | 0.67<br>[0.23, 1.98] | 1.21<br>[0.86, 1.71] | 1.07<br>[0.90, 1.28] | 0.78**<br>[0.62, 0.96] |
| **Reference: conceived post-rationing** |  |  |  |  |  |
| In-utero | 0.82*<br>[0.66, 1.02] | 0.72<br>[0.39, 1.34] | 0.93<br>[0.68, 1.26] | 0.99<br>[0.84, 1.18] | 0.83**<br>[0.70, 0.99] |
| In-utero + 6 months | 0.65***<br>[0.50, 0.86] | 0.42***<br>[0.23, 0.77] | 1.14<br>[0.82, 1.59] | 0.98<br>[0.84, 1.14] | 0.65***<br>[0.52, 0.81] |
| In-utero+ 12 months | 0.66**<br>[0.48, 0.92] | 0.22***<br>[0.11, 0.43] | 0.46***<br>[0.33, 0.64] | 0.77***<br>[0.63, 0.93] | 0.69***<br>[0.58, 0.84] |
| In-utero+18 months | 0.47***<br>[0.38, 0.59] | 0.18***<br>[0.08, 0.41] | 0.71**<br>[0.52, 0.98] | 0.84**<br>[0.72, 0.97] | 0.50***<br>[0.41, 0.61] |
| In-utero+24 months | 0.56***<br>[0.42, 0.76] | 0.32***<br>[0.14, 0.72] | 0.60***<br>[0.44, 0.82] | 0.64***<br>[0.53, 0.77] | 0.48***<br>[0.40, 0.59] |
| Observations | 62,858 | 62,858 | 62,858 | 35,298 | 27,560 |

*Notes:* Exponentiated coefficients from Cox proportional hazard models with clustered standard errors by month of birth. Hazard ratios (HR) reported with 95% confidence intervals in brackets.
\* p<0.10, \*\* p<0.05, \*\*\* p<0.01.

**Table S3:** Hazard ratios (95% CI) for cardiometabolic disease incidence by exposure duration to sugar rationing.

| Exposure duration<br>(Months) | Hypertension<br>(1) | Type 2<br>diabetes<br>(2) | Obesity<br>(3) | Heart<br>failure<br>(4) | Type 1<br>diabetes<br>(5) |
| --- | --- | --- | --- | --- | --- |
| –27 months | 1.02<br>[0.92, 1.13] | 1.00<br>[0.80, 1.26] | 1.10<br>[0.96, 1.26] | 1.12<br>[0.81, 1.56] | 1.42**<br>[1.07, 1.88] |
| –21 months | 1.02<br>[0.96, 1.09] | 0.98<br>[0.89, 1.09] | 1.05<br>[0.96, 1.15] | 1.16**<br>[1.01, 1.32] | 0.93<br>[0.75, 1.16] |
| –15 months | 1.02<br>[0.94, 1.10] | 0.95<br>[0.84, 1.08] | 1.00<br>[0.83, 1.21] | 0.89*<br>[0.79, 1.00] | 0.86<br>[0.64, 1.15] |
| **Reference: conceived post-rationing** |  |  |  |  |  |
| In-utero | 0.97<br>[0.92, 1.02] | 0.92<br>[0.83, 1.02] | 0.94<br>[0.83, 1.07] | 0.90*<br>[0.79, 1.02] | 1.02<br>[0.83, 1.26] |
| In-utero + 6 months | 0.97<br>[0.92, 1.02] | 0.87<br>[0.73, 1.03] | 0.91**<br>[0.83, 1.00] | 0.98<br>[0.88, 1.09] | 1.08<br>[0.83, 1.40] |
| In-utero + 12 months | 0.86***<br>[0.82, 0.91] | 0.79***<br>[0.68, 0.92] | 0.89*<br>[0.78, 1.00] | 0.81***<br>[0.69, 0.94] | 0.82*<br>[0.67, 1.00] |
| In-utero + 18 months | 0.92***<br>[0.87, 0.97] | 0.75***<br>[0.66, 0.85] | 0.79***<br>[0.71, 0.87] | 0.83**<br>[0.72, 0.96] | 0.88<br>[0.68, 1.15] |
| In-utero + 24 months | 0.84***<br>[0.79, 0.88] | 0.72***<br>[0.65, 0.79] | 0.78***<br>[0.70, 0.87] | 0.81***<br>[0.73, 0.89] | 0.82<br>[0.60, 1.12] |
| Observations | 62,686 | 62,686 | 62,686 | 62,686 | 62,686 |

*Notes:* Exponentiated coefficients from Cox proportional hazard models with clustered standard errors by month of birth. Hazard ratios (HR) reported with 95% confidence intervals in brackets.
\* p<0.10, \*\* p<0.05, \*\*\* p<0.01.

**Table S4:**
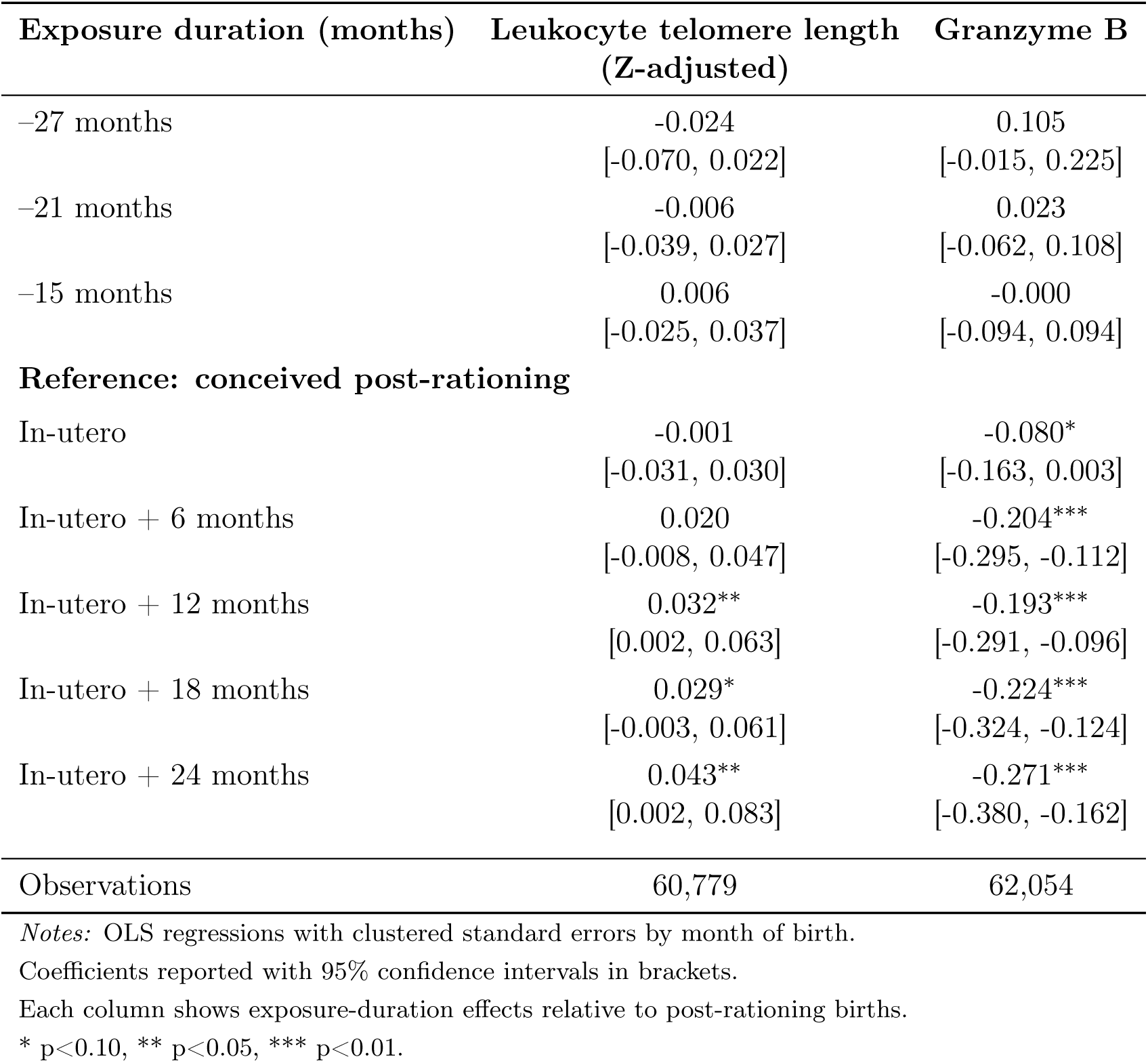
Coefficient estimates (95% CI) for biological ageing markers by exposure duration to sugar rationing.

**Table S5:** Coefficient estimates (95% CI) for nutrient intake by exposure duration to sugar rationing.

| Exposure duration<br>(months) | Food weight<br>(1) | Healthy eating<br>index<br>(2) | Food diversity<br>score<br>(3) |
| --- | --- | --- | --- |
| -27 months | 17.244<br>[-40.444, 74.933] | -0.043<br>[-0.898, 0.811] | -0.096<br>[-0.215, 0.024] |
| -21 months | 17.452<br>[-23.418, 58.323] | -0.139<br>[-0.744, 0.466] | -0.085*<br>[-0.170, -0.001] |
| -15 months | 5.320<br>[-40.215, 50.854] | 0.202<br>[-0.472, 0.877] | -0.005<br>[-0.100, 0.089] |
| **Reference: conceived post-rationing** |  |  |  |
| In-utero | -25.960<br>[-66.306, 14.385] | 0.692*<br>[0.094, 1.290] | 0.136**<br>[0.053, 0.220] |
| In-utero + 6 months | -41.197<br>[-84.869, 2.475] | 1.059**<br>[0.412, 1.706] | 0.244***<br>[0.153, 0.334] |
| In-utero + 12 months | -47.222*<br>[-94.255, -0.190] | 1.334***<br>[0.637, 2.031] | 0.270***<br>[0.172, 0.368] |
| In-utero + 18 months | -55.680*<br>[-104.152, -7.207] | 1.359***<br>[0.641, 2.077] | 0.333***<br>[0.232, 0.433] |
| In-utero + 24 months | -74.219**<br>[-126.661, -21.778] | 1.695***<br>[0.918, 2.472] | 0.377***<br>[0.268, 0.485] |
| Observations | 28,847 | 28,803 | 28,803 |

*Notes:* OLS regressions with clustered standard errors by month of birth.
Coefficients reported with 95% confidence intervals in brackets.
Each column shows exposure-duration effects relative to post-rationing births.
\* p<0.10, \*\* p<0.05, \*\*\* p<0.01.

**Table S6:** Coefficient estimates (95% CI) for nutrient intake by exposure duration to sugar rationing.

| Exposure duration<br>(months) | Sugars<br>(1) | Fibre<br>(2) | Starch<br>(3) | Fat<br>(4) | Protein<br>(5) |
| --- | --- | --- | --- | --- | --- |
| -27 months | -1.949<br>[-5.581, 1.684] | -0.099<br>[-0.580, 0.382] | -0.262<br>[-3.564, 3.039] | 0.000<br>[-2.193, 2.194] | -0.312<br>[-2.147, 1.523] |
| -21 months | -1.187<br>[-3.761, 1.387] | -0.009<br>[-0.350, 0.332] | 0.650<br>[-1.689, 2.989] | -0.658<br>[-2.212, 0.896] | 0.493<br>[-0.807, 1.794] |
| -15 months | -2.468<br>[-5.335, 0.399] | 0.042<br>[-0.338, 0.422] | -0.797<br>[-3.403, 1.809] | -0.368<br>[-2.099, 1.364] | -0.566<br>[-2.015, 0.882] |
| **Reference: conceived post-rationing** |  |  |  |  |  |
| In-utero | -3.752**<br>[-6.293, -1.211] | -0.131<br>[-0.467, 0.206] | -2.620*<br>[-4.929, -0.311] | -1.344<br>[-2.878, 0.190] | -1.125<br>[-2.409, 0.158] |
| In-utero+6 months | -2.263<br>[-5.013, 0.487] | 0.195<br>[-0.170, 0.559] | -0.592<br>[-3.092, 1.907] | -0.499<br>[-2.159, 1.162] | -0.036<br>[-1.425, 1.353] |
| In-utero+12 months | -4.108**<br>[-7.069, -1.146] | -0.084<br>[-0.476, 0.308] | -1.527<br>[-4.219, 1.164] | -0.405<br>[-2.193, 1.383] | -0.832<br>[-2.329, 0.664] |
| In-utero+18 months | -2.686<br>[-5.739, 0.366] | 0.142<br>[-0.262, 0.546] | -0.833<br>[-3.607, 1.942] | 0.253<br>[-1.590, 2.096] | -0.570<br>[-2.112, 0.972] |
| In-utero+24 months | -3.274*<br>[-6.576, 0.028] | 0.146<br>[-0.291, 0.584] | -1.113<br>[-4.114, 1.889] | 0.498<br>[-1.496, 2.492] | -0.230<br>[-1.898, 1.438] |
| Observations | 28,847 | 28,847 | 28,847 | 28,847 | 28,847 |

*Notes:* OLS regressions with clustered standard errors by month of birth.
Coefficients reported with 95% confidence intervals in brackets.
Each column shows exposure-duration effects relative to post-rationing births.
\* p<0.10, \*\* p<0.05, \*\*\* p<0.01.

**Table S7:** Sample distribution by birth year-month and status to sugar rationing (N = 64,761).

| Birth year-month | N | Percent | Rationing Status | Rationing Length |
| --- | --- | --- | --- | --- |
| 1951m10 | 1,166 | 1.80 | Rationed | Up to 24 months + in-utero |
| 1951m11 | 1,140 | 1.76 | Rationed | Up to 24 months + in-utero |
| 1951m12 | 1,245 | 1.92 | Rationed | Up to 24 months + in-utero |
| 1952m1 | 1,268 | 1.96 | Rationed | Up to 24 months + in-utero |
| 1952m2 | 1,248 | 1.93 | Rationed | Up to 24 months + in-utero |
| 1952m3 | 1,421 | 2.19 | Rationed | Up to 24 months + in-utero |
| 1952m4 | 1,286 | 1.99 | Rationed | Up to 18 months + in-utero |
| 1952m5 | 1,441 | 2.23 | Rationed | Up to 18 months + in-utero |
| 1952m6 | 1,324 | 2.04 | Rationed | Up to 18 months + in-utero |
| 1952m7 | 1,276 | 1.97 | Rationed | Up to 18 months + in-utero |
| 1952m8 | 1,223 | 1.89 | Rationed | Up to 18 months + in-utero |
| 1952m9 | 1,217 | 1.88 | Rationed | Up to 18 months + in-utero |
| 1952m10 | 1,220 | 1.88 | Rationed | Up to 12 months + in-utero |
| 1952m11 | 1,103 | 1.70 | Rationed | Up to 12 months + in-utero |
| 1952m12 | 1,190 | 1.84 | Rationed | Up to 12 months + in-utero |
| 1953m1 | 1,243 | 1.92 | Rationed | Up to 12 months + in-utero |
| 1953m2 | 1,237 | 1.91 | Rationed | Up to 12 months + in-utero |
| 1953m3 | 1,318 | 2.04 | Rationed | Up to 12 months + in-utero |
| 1953m4 | 1,338 | 2.07 | Rationed | Up to 6 months + in-utero |
| 1953m5 | 1,396 | 2.16 | Rationed | Up to 6 months + in-utero |
| 1953m6 | 1,281 | 1.98 | Rationed | Up to 6 months + in-utero |
| 1953m7 | 1,253 | 1.93 | Rationed | Up to 6 months + in-utero |
| 1953m8 | 1,235 | 1.91 | Rationed | Up to 6 months + in-utero |
| 1953m9 | 1,215 | 1.88 | Rationed | Up to 6 months + in-utero |
| 1953m10 | 1,131 | 1.75 | Rationed | In-utero only |
| 1953m11 | 1,097 | 1.69 | Rationed | In-utero only |
| 1953m12 | 1,084 | 1.67 | Rationed | In-utero only |
| 1954m1 | 1,186 | 1.83 | Rationed | In-utero only |
| 1954m2 | 1,179 | 1.82 | Rationed | In-utero only |
| 1954m3 | 1,282 | 1.98 | Rationed | In-utero only |
| 1954m4 | 1,178 | 1.82 | Rationed | In-utero only |
| 1954m5 | 1,350 | 2.08 | Rationed | In-utero only |
| 1954m6 | 1,208 | 1.87 | Rationed | In-utero only |
| 1954m7 | 1,204 | 1.86 | No rationing | - |
| 1954m8 | 1,205 | 1.86 | No rationing | - |
| 1954m9 | 1,098 | 1.70 | No rationing | - |
| 1954m10 | 1,118 | 1.73 | No rationing | - |
| 1954m11 | 1,036 | 1.60 | No rationing | - |
| 1954m12 | 1,096 | 1.69 | No rationing | - |
| 1955m1 | 1,123 | 1.73 | No rationing | - |
| 1955m2 | 1,082 | 1.67 | No rationing | - |
| 1955m3 | 1,230 | 1.90 | No rationing | - |
| 1955m4 | 1,160 | 1.79 | No rationing | - |
| 1955m5 | 1,273 | 1.97 | No rationing | - |
| 1955m6 | 1,194 | 1.84 | No rationing | - |
| 1955m7 | 1,180 | 1.82 | No rationing | - |
| 1955m8 | 1,049 | 1.62 | No rationing | - |
| 1955m9 | 1,046 | 1.62 | No rationing | - |
| 1955m10 | 1,095 | 1.69 | No rationing | - |
| 1955m11 | 1,046 | 1.62 | No rationing | - |
| 1955m12 | 1,105 | 1.71 | No rationing | - |
| 1956m1 | 1,097 | 1.69 | No rationing | - |
| 1956m2 | 1,069 | 1.65 | No rationing | - |
| 1956m3 | 1,276 | 1.97 | No rationing | - |

**Table S8:** Log-rank test results for equality of survival functions.

| **Disease outcome** | **Chi-square statistic** | **p-value** |
| --- | --- | --- |
| Liver cancer | 14.59 | < 0.001 |
| Rectum cancer | 9.01 | 0.003 |
| Lung cancer | 23.63 | < 0.001 |
| Breast cancer | 36.54 | < 0.001 |
| Prostate cancer | 24.89 | < 0.001 |
| Hypertension | 36.01 | < 0.001 |
| Obesity | 12.70 | < 0.001 |
| Heart failure | 90.67 | < 0.001 |
| T2D | 24.09 | < 0.001 |
| T1D | 1.58 | 0.209 |

## Footnotes

1 However, randomized controlled trials in animals are possible and provide stronger causal evidence. Mice randomly assigned to high-sucrose or fructose diets exhibit higher tumour incidence and faster growth even in the absence of obesity. [10] show that sucrose promotes mammary tumour metastasis through inflammatory lipid pathways, while [11] find that modest fructose intake accelerates intestinal tumour growth by enhancing tumour metabolism. Nonetheless, a substantial translational gap remains between these controlled animal studies and human evidence, given marked differences in metabolic responses, exposure durations, and tumour biology across species [12, 13].

2 Butyrate is a vital energy source for colonocytes and has anti-carcinogenic properties; its depletion increases susceptibility to colorectal malignancies.

## References

[1] United Nations. Transforming our world: the 2030 Agenda for Sustainable Development; 2015. Accessed: 2025-11-16. Available from: https://sdgs.un.org/2030agenda.

[2] Avena NM, Rada P, Hoebel BG. Evidence for sugar addiction: behavioral and neurochem-ical effects of intermittent, excessive sugar intake. Neuroscience & biobehavioral reviews. 2008;32(1):20–39.

[3] Lustig RH, Schmidt LA, Brindis CD. Public health: The toxic truth about sugar. Nature. 2012;482:27–9.

[4] Pavlova NN, Thompson CB. The emerging hallmarks of cancer metabolism. Cell Metabolism. 2016;23(1):27–47.

[5] Pollak M. Insulin and insulin-like growth factor signalling in neoplasia. Nature Reviews Cancer. 2008;8(12):915–28.

[6] Coussens LM, Werb Z. Inflammation and cancer. Nature. 2002;420(6917):860–7.

[7] Makarem N, Lin Y, Bandera EV, Jacques PF, Parekh N. Consumption of sugar-sweetened beverages and cancer risk: results from the Framingham Offspring cohort (1971–2013). British Journal of Cancer. 2018;119(1):109–17.

[8] Fiolet T, Srour B, Sellem L, Kesse-Guyot E, Allès B, Méjean C, et al. Consumption of ultra-processed foods and cancer risk: results from NutriNet-Santé prospective cohort. bmj. 2018;360.

[9] Romaguera D, Norat T, Wark PA, Vergnaud AC, Schulze MB, van Woudenbergh GJ, et al. Consumption of sweet beverages and colorectal cancer risk in the European Prospective Investigation into Cancer and Nutrition (EPIC). British Journal of Nutrition. 2013;110(7):1353–62.

[10] Cao Y, Karin M, et al. Dietary sucrose promotes mammary tumor growth and metastasis in mice. Cancer Research. 2016;76(1):24–9.

[11] Gonçalves MD, Lu C, Tutnauer J, Hartman TE, Hwang SH, Murphy CJ, et al. High-fructose corn syrup enhances intestinal tumor growth in mice. Science. 2019;363(6433):1345–9.

[12] Nair AB, Jacob S. Translational research in cancer: challenges and opportunities. Oncology Reviews. 2015;9(1):283.

[13] van Norman GA. Limitations of animal studies for predicting toxicity in clinical trials: is it time to rethink our approach? JACC: Basic to Translational Science. 2019;4(7):845–54.

[14] Bray GA, Popkin BM. Dietary sugar and body weight: have we reached a crisis in the epidemic of obesity and diabetes? Health be damned! Pour on the sugar. Diabetes care. 2014;37(4):950–6.

[15] Yuan S, Kar S, Carter P, Vithayathil M, Mason AM, Burgess S, et al. Insulin, glucose, and cancer risk: a Mendelian randomisation study. Diabetologia. 2021;64:1132–44.

[16] Carreras-Torres R, Johansson M, Haycock PC, Relton C, Davey Smith G, Brennan P. The causal association between insulin resistance and cancer: a Mendelian randomization analysis. International Journal of Cancer. 2020;146(11):3023–30.

[17] Mohammadnejad S, Xu L, Zhou A, Haycock PC, Yarmolinsky J, Carter P, et al. Sugar intake, glycaemic traits, and breast cancer risk: a Mendelian randomisation study. BMC Medicine. 2023;21(1):94.

[18] Johnson RJ, Segal MS, Sautin Y, Nakagawa T, Feig DI, Kang DH, et al. Potential role of sugar (fructose) in the epidemic of hypertension, obesity and the metabolic syndrome, diabetes, kidney disease, and cardiovascular disease. American Journal of Clinical Nutrition. 2007;86(4):899–906.

[19] Jayedi A, Shab-Bidar S. Sugar-sweetened beverages and risk of cancer: an umbrella re-view of meta-analyses of cohort studies. Critical Reviews in Food Science and Nutrition. 2023;63(38):6397–411.

[20] Ripken DA, Franco OH, Voortman T. Dietary sugar intake and human health: a compre-hensive review. Nutrients. 2023;15(4):810.

[21] Barker DJ. The fetal and infant origins of adult disease. BMJ: British Medical Journal. 1990;301(6761):1111.

[22] Roseboom TJ, van der Meulen JHP, Ravelli ACJ, Osmond C, Barker DJP, Bleker OP. Effects of prenatal exposure to the Dutch famine on adult disease in later life: an overview. Molecular and Cellular Endocrinology. 2001;185(1–2):93–8.

[23] Jackson D. Caries experience in english children and young adults during the years 1947-1972. British dental journal. 1974;137(3):91–8.

[24] Gertler P, Gracner T. The sweet life: The long-term effects of a sugar-rich early childhood. National Bureau of Economic Research; 2022.

[25] Gracner T, Boone C, Gertler PJ. Exposure to sugar rationing in the first 1000 days of life protected against chronic disease. Science. 2024;386(6725):1043–8.

[26] WHO. Guideline: Sugars intake for adults and children. World Health Organization; 2015.

[27] Perez-Cornago A, Pollard Z, Young H, van Uden M, Andrews C, Piernas C, et al. Descrip-tion of the updated nutrition calculation of the Oxford WebQ questionnaire and comparison with the previous version among 207,144 participants in UK Biobank. European Journal of Clinical Nutrition. 2021;75(5):885–92.

[28] Birch LL. Development of food preferences. Annual Review of Nutrition. 1999;19:41–62.

[29] Mennella JA. Ontogeny of taste preferences: basic biology and implications for health. The American journal of clinical nutrition. 2014;99(3):704S–711S.

[30] Giovannucci E. Insulin, insulin-like growth factors and colon cancer: a review of the evi-dence. Journal of Nutrition. 2001;131(11 Suppl):3109S–3120S.

[31] Renehan AG, Tyson M, Egger M, Heller RF, Zwahlen M. Body-mass index and incidence of cancer: a systematic review and meta-analysis of prospective observational studies. The Lancet. 2008;371(9612):569–78.

[32] Zheng J, Zhou Z, Huang J, Tu Q, Wu H, Yang Q, et al. Exposure to sugar rationing in first 1000 days after conception and long term cardiovascular outcomes: natural experiment study. bmj. 2025;391.

[33] Mantovani A, Allavena P, Sica A, Balkwill F. Cancer-related inflammation. Nature. 2008;454(7203):436–44.

[34] Codd V, Denniff M, Swinfield C, Warner SC, et al. A major population resource of 474,074 participants in UK Biobank to investigate determinants and biomedical consequences of leukocyte telomere length. medRxiv. 2021. Preprint.

[35] Benetos A, Kark JD, Susser E, et al. Tracking and fixed ranking of leukocyte telomere length across the adult life course. Aging Cell. 2013;12(4):615–21.

[36] Demanelis K, Jasmine F, Chen LS, et al, . Determinants of telomere length across human tissues. Science. 2020;369(6509):eaaz6876.

[37] Department for Environment, Food and Rural Affairs. Family Food: Historic Reports, 1940–1984. National Food Survey historic dataset; 2016. Available from: https://www.gov.uk/government/statistics/family-food-historic-reports.

[38] Dewdney B, Roberts A, Qiao L, George J, Hebbard L. A sweet connection? Fructose’s role in hepatocellular carcinoma. Biomolecules. 2020;10(4):496.

[39] Giovannucci E. Insulin, insulin-like growth factors and colon cancer: a review of the evi-dence. The Journal of nutrition. 2001;131(11):3109S–3120S.

[40] Coussens LM, Werb Z. Inflammation and cancer. Nature. 2002;420(6917):860–7.

[41] Walker RW, Goran MI. Laboratory determined sugar content and composition of commer-cial infant formulas, baby foods and common grocery items targeted to children. Nutrients. 2015;7:5850–67.

[42] Cioffi CE, Figueroa J, Welsh JA. Added sugar intake among pregnant women in the United States: National Health and Nutrition Examination Survey 2003–2012. Journal of the Academy of Nutrition and Dietetics. 2018;118:886–95.e1.

[43] Snetselaar LG, de Jesus JM, DeSilva DM, Stoody EE. Dietary Guidelines for Americans, 2020–2025: Understanding the scientific process, guidelines, and key recommendations. Nutrition Today. 2021;56:287–95.

[44] Ratzmann KP, Steindel E, Hildebrandt R, Kohlhoff R. Is there a relationship between metabolic control and glucose concentration in breast milk of type 1 (insulin-dependent) diabetic mothers? Experimental and Clinical Endocrinology. 1988;92:32–6.

[45] Herrick KA, Fryar CD, Hamner HC, Park S, Ogden CL. Added sugars intake among US infants and toddlers. Journal of the Academy of Nutrition and Dietetics. 2020;120:23–32.

[46] Awad R, Kowash M, Hussein I, Salami A, Abdo M, Al-Halabi M. Sugar content in in-fant formula: Accuracy of labeling and conformity to guidelines. International Journal of Paediatric Dentistry. 2023;33:63–73.

[47] Ventura AK, Mennella JA. Innate and learned preferences for sweet taste during childhood. Current Opinion in Clinical Nutrition & Metabolic Care. 2011;14(4):379–84.

[48] Benton D. The influence of dietary status on the cognitive performance of children. Molecular nutrition & food research. 2010;54(4):457–70.

[49] Blackburn EH, Epel ES, Lin J. Human telomere biology: a contributory and interactive factor in aging, disease risks, and protection. Science. 2015;350(6265):1193–8.

[50] Epel ES, Blackburn EH, Lin J, Dhabhar FS, Adler NE, Morrow JD, et al. Accelerated telomere shortening in response to life stress. Proceedings of the National Academy of Sciences. 2004;101(49):17312–5.

[51] Wikby A, Nilsson BO, Forsey R, Thompson J, Strindhall J, Lofgren S, et al. The immune risk phenotype is associated with IL-6 in the terminal decline stage: The Swedish NONA immune longitudinal study. Mechanisms of Ageing and Development. 2005;126(11):1185–91.

[52] Chou JP, Ramirez CM, Wu JE, Effros RB. Accelerated aging in HIV/AIDS: Novel biomark-ers of senescent human CD8+ T cells. PLoS ONE. 2013;8(5):e64702.

[53] Rea IM, Gibson DS, McGilligan V, McNerlan SE, Alexander HD, Ross OA. Age and age-related diseases: Role of inflammation triggers and cytokines. Frontiers in Immunology. 2018;9:586.

[54] of Health UD, Care S. National Diet and Nutrition Survey: 2019 to 2023 Report; 2023. UK government nutrition statistics. Available from: https://www.gov.uk/government/statistics/national-diet-and-nutrition-survey-2019-to-2023.

[55] OECD, FAO. OECD–FAO Agricultural Outlook 2025–2034: Sugar; 2025. Global per-capita sugar consumption projections. Available from: https://www.oecd.org.

[56] Olovnikov AM. A theory of marginotomy: The incomplete copying of template margin in enzymic synthesis of polynucleotides and biological significance of the phenomenon. Journal of Theoretical Biology. 1973;41(1):181–90.

[57] O’Donovan A, Pantell MS, Puterman E, Dhabhar FS, Blackburn EH, Yaffe K, et al. Cellular and molecular mechanisms of stress-related immune dysregulation and relevance to disease. Brain, Behavior, and Immunity. 2011;25(1):19–28.

[58] Epel ES. Psychological and metabolic stress: a recipe for accelerated cellular aging? Hor-mones. 2009;8(1):7–22.

[59] von Zglinicki T. Oxidative stress shortens telomeres. Trends in Biochemical Sciences. 2002;27(7):339–44.

[60] Müezzinler A, Zaineddin AK, Brenner H. Telomere length and mortality in adults: A systematic review and meta-analysis. Ageing Research Reviews. 2013;12(2):509–19.

[61] Wentzensen N, Mirabello L, Pfeiffer RM, Savage SA. Leukocyte telomere length in re-lation to cancer risk: A prospective study of 47,102 US men and women. PLoS One. 2011;6(2):e16918.

[62] Willeit P, Willeit J, Mayr A, Weger S, Oberhollenzer F, Brandstätter A, et al. Telomere length and risk of incident cancer and cancer mortality. JAMA. 2010;304(1):69–75.

[63] Wang Q, Zhan Y, Pedersen NL, Fang F, Hägg S. Telomere length and all-cause mortality: A meta-analysis. Ageing Research Reviews. 2018;48:11–20.

[64] Zhan Y, Song C, Karlsson R, Tillander A, Reynolds CA, Pedersen NL, et al. Telomere length shortening and Alzheimer’s disease: A Mendelian randomization study. Journal of Alzheimer’s Disease. 2015;46(2):619–28.

[65] Codd V, Nelson CP, Albrecht E, Mangino M, Deelen J, Buxton JL, et al. Identification of seven loci affecting mean telomere length and their association with disease. Nature Genetics. 2013;45(4):422–7.

[66] Kennedy ET, Ohls J, Carlson A, Fleming K. The Healthy Eating Index: Design and applications. Journal of the American Dietetic Association. 1995;95(10):1103–8.

[67] Guenther PM, Reedy J, Krebs-Smith SM, Reeve BB. Development and evaluation of the Healthy Eating Index-2005: Technical report. Center for Nutrition Policy and Promotion, US Department of Agriculture. 2008.

[68] Kant AK, Schatzkin A, Graubard BI, Schairer C. A prospective study of diet quality and mortality in women. JAMA. 2000;283(16):2109–15.

[69] Wiafe MA, Apprey C, Annan RA. Dietary diversity and nutritional status of adolescents in rural Ghana. Nutrition and Metabolic Insights. 2023;16:11786388231158487.

[70] Zweiniger-Bargielowska I. Austerity in Britain: Rationing, Controls, and Consumption, 1939–1955. Oxford: Oxford University Press; 2000.

[71] Toverud G. The influence of war and post-war conditions on the teeth of Norwegian school children. Journal of the American Dental Association. 1949;38(6):691–706.

[72] Hollingsworth DF. Dental caries in Britain, 1939–1946. British Dental Journal. 1974;137(1):23–28.

[73] Hollingsworth DF. Changes in dental health during and after the Second World War. Community Dental Health. 1983;1(2):95–102.

[74] Prynne CJ, Paul AA, Price GM, Day KC, Hilder WS, Wadsworth MEJ. Food and nutrient intake of a national sample of 4-year-old children in 1950: comparison with the 1990s. Public Health Nutrition. 1999;2(4):537–547.

